# Model-informed COVID-19 vaccine prioritization strategies by age and serostatus

**DOI:** 10.1101/2020.09.08.20190629

**Authors:** Kate M. Bubar, Kyle Reinholt, Stephen M. Kissler, Marc Lipsitch, Sarah Cobey, Yonatan H. Grad, Daniel B. Larremore

**Affiliations:** Department of Applied Mathematics, University of Colorado Boulder, Boulder, CO, 80303, USA; IQ Biology Program, University of Colorado Boulder, Boulder, CO, 80309, USA; Department of Computer Science, University of Colorado Boulder, Boulder, CO, 80309, USA; Department of Immunology and Infectious Diseases, Harvard T.H. Chan School of Public Health, Boston, MA, 02115, USA; Center for Communicable Disease Dynamics, Harvard T.H. Chan School of Public Health, Boston, MA, 02115, USA; Department of Ecology and Evolution, University of Chicago, Chicago, IL, 60637, USA; BioFrontiers Institute, University of Colorado Boulder, Boulder, CO, 80303, USA

## Abstract

Limited initial supply of SARS-CoV-2 vaccine raises the question of how to prioritize available doses. Here, we used a mathematical model to compare five age-stratified prioritization strategies. A highly effective transmission-blocking vaccine prioritized to adults ages 20-49 years minimized cumulative incidence, but mortality and years of life lost were minimized in most scenarios when the vaccine was prioritized to adults over 60 years old. Use of individual-level serological tests to redirect doses to seronegative individuals improved the marginal impact of each dose while potentially reducing existing inequities in COVID-19 impact. While maximum impact prioritization strategies were broadly consistent across countries, transmission rates, vaccination rollout speeds, and estimates of naturally acquired immunity, this framework can be used to compare impacts of prioritization strategies across contexts.

---

SARS-CoV-2 has caused a public health and economic crisis worldwide. As of January 2021, there have been over 85 million cases and 1.8 million deaths reported (*1*). To combat this crisis, a variety of non-pharmaceutical interventions have been implemented, including shelter-in-place orders, limited travel, and remote schooling. While these efforts are essential to slowing transmission in the short term, long-term solutions—such as vaccines that protect from SARS-CoV-2 infection—remain urgently needed. The benefits of an effective vaccine for individuals and their communities have resulted in widespread demand, so it is critical that decision-making on vaccine distribution is well motivated, particularly in the initial phases when vaccine availability is limited (*2*).

Here, we employ a model-informed approach to quantify the impact of COVID-19 vaccine prioritization strategies on cumulative incidence, mortality, and years of life lost. Our approach explicitly addresses variation in three areas that can influence the outcome of vaccine distribution decisions. First, we consider variation in the performance of the vaccine, including its overall efficacy, a hypothetical decrease in efficacy by age, and the vaccine’s ability to block transmission. Second, we consider variation in both susceptibility to infection and the infection fatality rate by age. Third, we consider variation in the population and policy, including the age distribution, age-stratified contact rates, and initial fraction of seropositive individuals by age, and the speed and timing of the vaccine’s rollout relative to transmission. While the earliest doses of vaccines will be given to front-line health care workers under plans such as those from the COVAX initiative and the US NASEM recommendations (*3*), our work is focused on informing the prioritization of the doses that follow. Based on regulatory approvals and initial vaccine rollout speeds of early 2021, our investigation focuses generally on scenarios with a partially mitigated pandemic (*R* between 1.1 and 2.0), vaccines with protective efficacy of 90%, and rollout speeds of 0.2% of the population per day.

There are two main approaches to vaccine prioritization: (1) directly vaccinate those at highest risk for severe outcomes and (2) protect them indirectly by vaccinating those who do the most transmitting. Model-based investigations of the tradeoffs between these strategies for influenza vaccination have led to recommendations that children be vaccinated due to their critical role in transmission (*4, 5*) and have shown that direct protection is superior when reproduction numbers are high but indirect protection is superior when transmission is low (*6*). Similar modeling for COVID-19 vaccination has found that the optimal balance between direct and indirect protection depends on both vaccine efficacy and supply, recommending direct vaccination of older adults for low-efficacy vaccines and for high-efficacy but supply-limited vaccines (*7*). Rather than comparing prioritization strategies, others have compared hypothetical vaccines, showing that even those with lower efficacy for direct protection may be more valuable if they also provide better indirect protection by blocking transmission (*8*). Prioritization of transmission-blocking vaccines can also be dynamically updated based on the current state of the epidemic, shifting prioritization to avoid decreasing marginal returns (*9*). These efforts to prioritize and optimize doses complement other work showing that, under different vaccine efficacy and durability of immunity, the economic and health benefits of COVID-19 vaccines will be large in the short and medium terms (*10*). The problem of vaccine prioritization also parallels the more general problem of optimal resource allocation to reduce transmission, e.g. with masks (*11*).

## Evaluation of vaccine prioritization strategies

We evaluated the impact of vaccine prioritization strategies using an age-stratified SEIR model, because age has been shown to be an important correlate of susceptibility (*12–14*), seroprevalence (*12, 15*), severity (*16–18*), and mortality (*19, 20*). This model includes an age-dependent contact matrix, susceptibility to infection, and infection fatality rate (IFR), allowing us to estimate cumulative incidence of SARS-CoV-2 infections, mortality due to infection, and years of life lost (YLL; see Methods) via forward simulations of one year of disease dynamics. Cumulative incidence, mortality, and YLL were then used as outcomes by which to compare vaccine prioritization strategies. These comparisons may be explored using accompanying open-source and interactive calculation tools that accompany this study (*21*).

We first examined the impact of five vaccine prioritization strategies for a hypothetical infection- and transmission-blocking vaccine of varying efficacy. The strategies prioritized vaccines to (1) children and teenagers, (2) adults between ages 20 and 49 years, (3) adults 20 years or older, (4) adults 60 years or older, and (5) all individuals (Fig. 1A). In all strategies, once the prioritized population was vaccinated, vaccines were allocated irrespective of age, i.e. in proportion to their numbers in the population. To incorporate vaccine hesitancy, at most 70% of any age group was eligible to be vaccinated (*24*).

**Figure 1:**
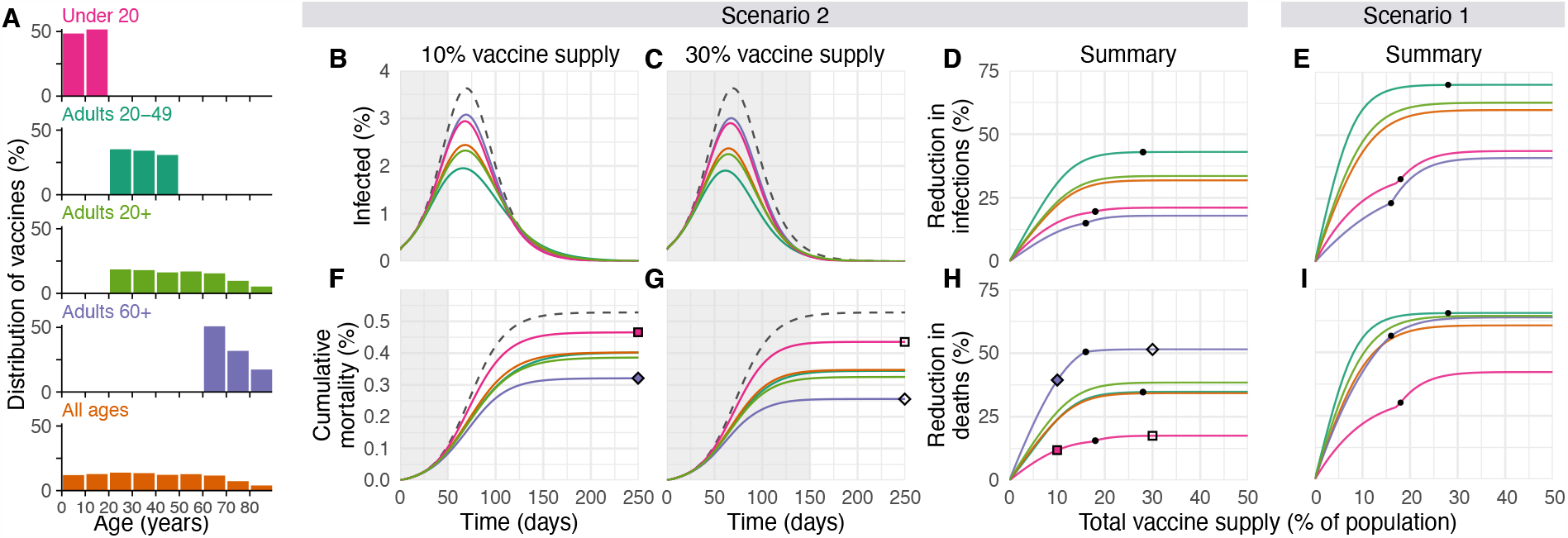
Impacts of vaccine prioritization strategies on mortality and infections. (A) Distribution of vaccines for five prioritization strategies: under 20, adults 20-49, adults 20+, adults 60+ and all ages. (B, C) Example simulation curves show percentage of the total population infected over time and (F, G) cumulative mortality for no vaccines (grey dashed lines) and for five different prioritization strategies (colored lines matching panel A), with 10% (B, F) and 30% (C, G) vaccine supply. Summary curves show percent reductions in (D, E) infections and (H, I) deaths in comparison to an unmitigated outbreak for vaccine supplies between 1% and 50% after 365 days of simulation. Squares and diamonds show how the outputs from single simulations (F, G) correspond to points in summary curves (H). Grey shading indicates period during which vaccine is being rolled out at 0.2% of total population per day. Black dots indicate breakpoints at which prioritized demographic groups have been 70% vaccinated, after which vaccines are distributed without prioritization. These simulations assume contact patterns and demographics of the United States (*22, 23*) and an all-or-nothing, transmission-blocking vaccine with 90% vaccine efficacy and *R*_0_ = 1.5 (Scenario 2) and *R*_0_ = 1.15 (Scenario 1).

We measured reductions in cumulative incidence, mortality, and YLL achieved by each strategy, varying the vaccine supply between 1% and 50% of the total population, under two scenarios. In Scenario 1, vaccines were administered to 0.2% of the population per day until supply was exhausted, with *R*_0_ = 1.15, representing highly mitigated spread during vaccine rollout. In Scenario 2, vaccines were administered to 0.2% of the population per day until supply was exhausted, but with *R*_0_ = 1.5, representing substantial viral growth during vaccine rollout (see Fig. 1 for example model outputs). Results for additional scenarios in which vaccines were administered before transmission began are described in Supplementary Text, corresponding to countries without ongoing community spread such as South Korea and New Zealand. We considered two ways in which vaccine efficacy (*ve*) could be below 100%: an all-or-nothing vaccine, where the vaccine provides perfect protection to a fraction *ve* of individuals who receive it, or as a leaky vaccine, where all vaccinated individuals have reduced probability *ve* of infection after vaccination (see Methods).

Of the five strategies, direct vaccination of adults over 60 years (60+) always reduced mortality and YLL more than the alternative strategies when transmission was high (*R*_0_ = 1.5; Scenario 2; 90% efficacy, Fig. 1; 30%-100% efficacy, Fig. S5). For lower transmission (*R*_0_ = 1.15; Scenario 1), vaccination of adults 20-49 reduced mortality and YLL more than the alternative strategies, but differences between prioritization of adults 20-49, adults 20+, and adults 60+ were small for vaccine supplies above 25% (Figs. 1 and S5). Prioritizing adults 20-49 minimized cumulative incidence in both scenarios for all vaccine efficacies (Figs. 1 and S5). Prioritizing adults 20-49 also minimized cumulative incidence in both scenarios under alternative rollout speeds (0.05% to 1% vaccinated per day; Fig. S6). When rollout speeds were at least 0.3% per day and vaccine supply covered at least 25% of the population, the mortality minimizing strategy shifted from prioritization of ages 20-49 to adults 20+ or adults 60+ for Scenario 1; when rollout speeds were at least 0.75% per day and covered at least 24% of the population, the mortality minimizing strategy shifted from prioritization of adults 60+ to adults 20+ or 20-49 for Scenario 2 (Fig. S6). Findings for mortality and YLL were only slightly changed by modeling vaccine efficacy as all-or-nothing (Fig. S5) or leaky (Fig. S7).

### Impact of transmission rates, age demographics, and contact structure

To evaluate the impact of transmission rates on the strategy that most reduced mortality, we varied the basic reproductive number *R*_0_ from 1.1 to 2.0 when considering a hypothetical infection- and transmission-blocking vaccine with 90% vaccine efficacy. We found that prioritizing adults 60+ remained the best way to reduce mortality and YLL for *R*_0_ *≥*1.3, but prioritizing adults 20-49 was superior for *R*_0_ *≤* 1.2 (Fig. 2A, B and S8). Prioritizing adults 20-49 minimized infections for all values of *R*_0_ investigated (Fig. S8).

**Figure 2:**
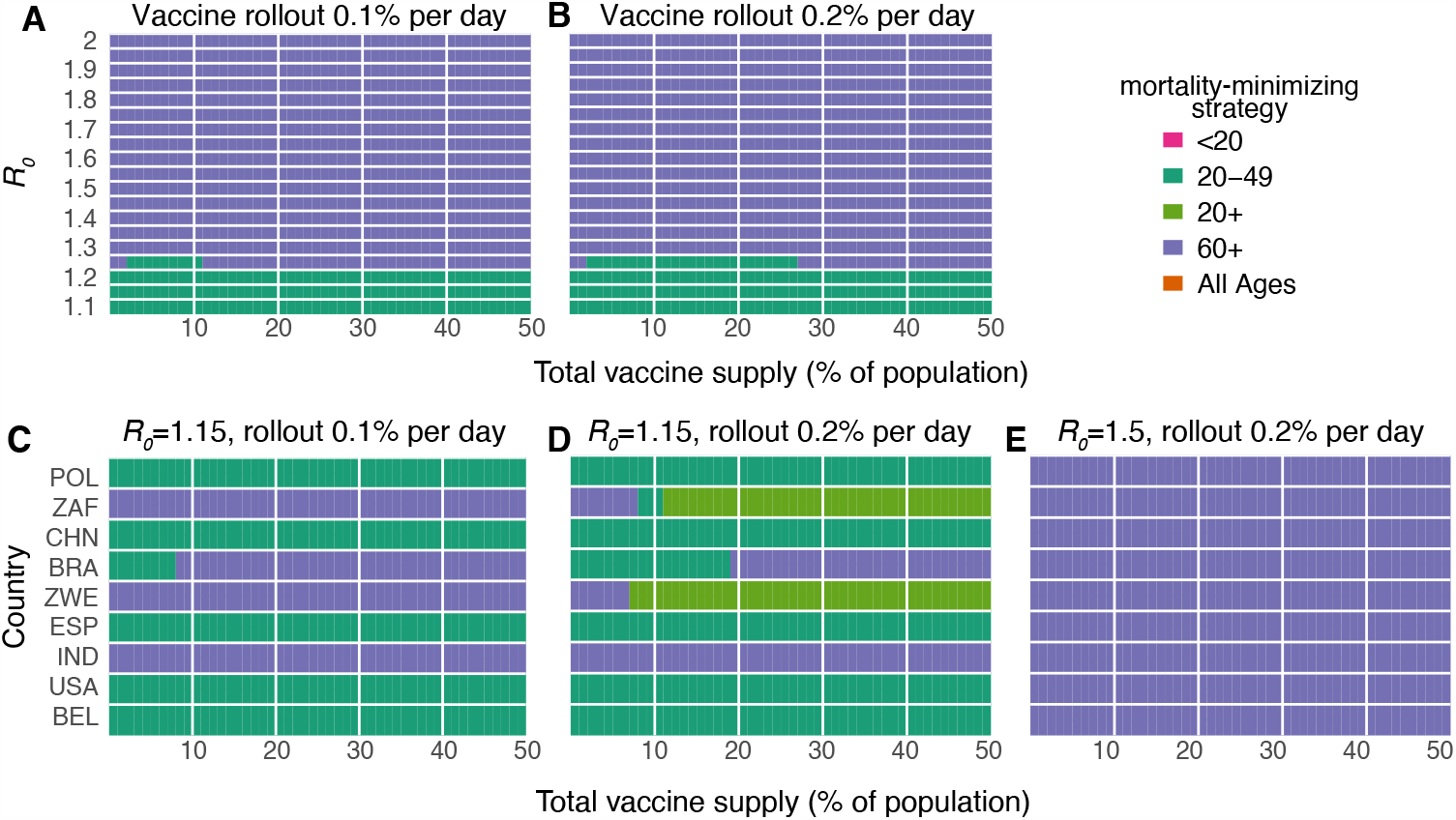
Mortality-minimizing vaccine prioritization strategies across reproductive numbers R_0_ and countries. Heatmaps show the prioritization strategies resulting in maximum reduction of mortality for varying values of the basic reproductive number *R*_0_ (A, B) and across nine countries (C, D, E), for vaccine supplies between 1% and 50% of the total population, for an all-or-nothing and transmission blocking vaccine, 90% vaccine efficacy. (A, B) Shown: contact patterns and demographics of the United States (*22,23*); (C, D, E) Shown: contact patterns and demographics of POL, Poland; ZAF, South Africa; CHN, China; BRA, Brazil; ZWE, Zimbabwe; ESP, Spain; IND, India; USA, United States of America; BEL, Belgium, with *R*_0_ and rollout speeds as indicated.

To determine whether our findings were robust across countries, we analyzed the ranking of prioritization strategies for populations with the age distributions and modeled contact structures of the United States, Belgium, Brazil, China, India, Poland, South Africa, and Spain. Across these countries, direct vaccination of adults 60+ minimized mortality for all levels of vaccine supply when transmission was high (*R*_0_ = 1.5, Scenario 2; Fig. 2E), but in only some cases when transmission was lower (*R*_0_ = 1.15, rollout 0.2% per day, Scenario 1; Fig. 2D). Decreasing rollout speed from 0.2% to 0.1% per day caused prioritization of adults 60+ to be favored in additional scenarios (Fig. 2C). Across countries, vaccination of adults 20-49 nearly always minimized infections, and vaccination of adults 60+ nearly always minimized YLL for Scenario 2, but no clear ranking of strategies emerged consistently to minimize YLL in Scenario 1 (Fig. S9).

### Vaccines with imperfect transmission blocking effects

We also considered whether the rankings of prioritization strategies to minimize mortality would change if a vaccine were to block COVID-19 symptoms and mortality with 90% efficacy but with variable impact on SARS-CoV-2 infection and transmission. We found that direct vaccination of adults 60+ minimized mortality for all vaccine supplies and transmission-blocking effects under Scenario 2, and for all vaccine supplies when up to 50% of transmission was blocked in Scenario 1 (Supplementary Text and Supplementary Fig. S10).

### Variation in vaccine efficacy by age

COVID-19 vaccines may not be equally effective across age groups in preventing infection or disease, a phenomenon known to affect influenza vaccines (*25–28*). To understand the impact of age-dependent COVID-19 vaccine efficacy, we incorporated a hypothetical linear decrease from a baseline efficacy of 90% for those under 60 to 50% in those 80 and older (Fig. 3). As expected, this diminished the benefits of any prioritization strategy that included older adults. For instance, strategies prioritizing adults 20-49 were unaffected by decreased efficacy among adults 60+, while strategies prioritizing adults 60+ were markedly diminished (Fig. 3). Despite these effects, prioritization of adults 60+ remained superior to the alternative strategies to minimize mortality in Scenario 2.

**Figure 3:**
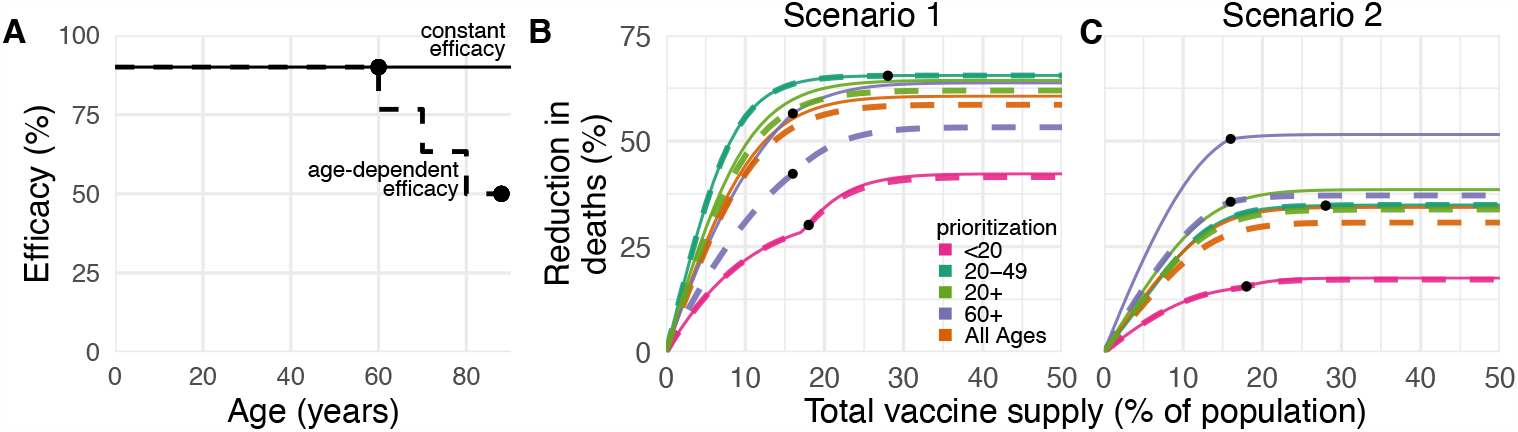
Effects of age-dependent vaccine efficacy on the impacts of prioritization strategies. (A) Diagram of hypothetical age-dependent vaccine efficacy shows decrease from 90% baseline efficacy to 50% efficacy among individuals 80+ beginning at age 60 (dashed line). (B, C) Percent reduction in deaths in comparison to an unmitigated outbreak for transmission-blocking all-or-nothing vaccines with either constant 90% efficacy for all age groups (solid lines) or age-dependent efficacy shown in panel A (dashed lines), covering Scenario 1 (0.2% rollout/day, *R*_0_ = 1.15; B) and Scenario 2 (0.2% rollout/day, *R*_0_ = 1.5; C). Black dots indicate breakpoints at which prioritized demographic groups have been 70% vaccinated, after which vaccines are distributed without prioritization. Shown: contact patterns and demographics of the United States (*22, 23*); all-or nothing and transmission blocking vaccine.

To test whether more substantial age-dependent vaccine effects would change which strategy minimized mortality in Scenario 2, we varied the onset age of age-dependent decreases in efficacy, the extent to which it decreased, and the baseline efficacy from which it decreased. We found that as long as the age at which efficacy began to decrease was 70 or older and vaccine efficacy among adults 80+ was at least 25%, prioritizing adults 60+ remained superior in the majority of parameter combinations. This finding was robust to whether the vaccine was modeled as leaky vs all-or-nothing, but we observed considerable variation from country to country (Fig. S11).

### Incorporation of population seroprevalence and individual serological testing

Due to early indications that naturally acquired antibodies correlate with protection from reinfection (*29*), seroprevalence will affect vaccine prioritization in two ways. First, depending on the magnitude and age distribution of seroprevalence at the time of vaccine distribution, the ranking of strategies could change. Second, distributing vaccines to seropositive individuals would reduce the marginal benefit of vaccination per dose.

To investigate the impact of vaccinating mid-epidemic while using serology to target the vaccine to seronegative individuals, we included age-stratified seroprevalence estimates in our model by moving the data-specified proportion of seropositive individuals from susceptible to recovered status. We then simulated two approaches to vaccine distribution. In the first, vaccines were distributed according to the five prioritization strategies introduced above, regardless of any individual’s serostatus. In the second, vaccines were distributed with a serological test, such that individuals with a positive serological test would not be vaccinated, allowing their dose to be given to someone else in their age group.

We included age-stratified seroprevalence estimates from New York City [August 2020; overall sero-prevalence 26.9% (*30*)] and demographics and age-contact structure from the United States in evaluations of the previous five prioritization strategies. For this analysis, we focused on Scenario 2 (0.2% rollout per day, *R* = 1.5 inclusive of seropositives), and found that the ranking of strategies to minimize incidence, mortality, and YLL remained unchanged: prioritizing adults 60+ most reduced mortality and prioritizing adults 20-49 most reduced incidence, regardless of whether vaccination was limited to seronegative individuals (Fig. 4). These rankings were unchanged when we used lower or higher age-stratified seroprevalence estimates to test the consistency of results (Connecticut, July 2020, overall seroprevalence 3.4% (*31*) and synthetic, overall seroprevalence 39.5%; Figs. S12 and S13). Despite lowered sensitivity to detect past exposure due to seroreversion (*32, 33*), preferentially vaccinating seronegative individuals yielded large additional reductions in cumulative incidence and mortality in locations with higher seroprevalence (Figs. 4 and S13) and modest reductions in locations with low seroprevalence (Fig. S12). These results remained unchanged when statistical uncertainty, due to sample size and imperfect test sensitivity and specificity, were incorporated into the model (*34*).

**Figure 4:**
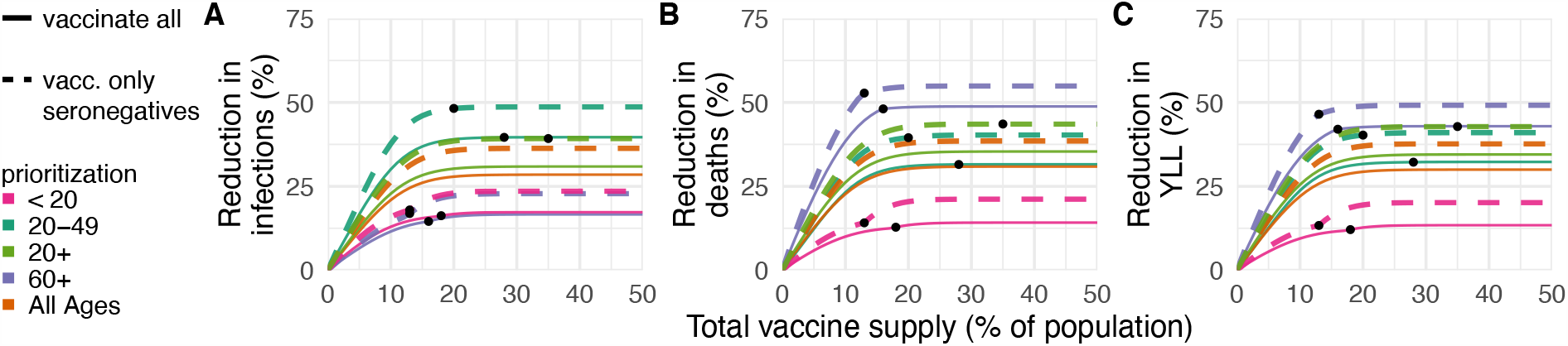
Effects of existing seropositivity on the impacts of prioritization strategies. Percent reductions in (A) infections, (B) deaths, and (C) years of life lost (YLL) for prioritization strategies when existing age-stratified seroprevalence is incorporated [August 2020 estimates for New York City; mean seroprevalence 26.9% (*30*)]. Plots show reductions for Scenario 2 (0.2% rollout/day, *R* = 1.5) when vaccines are given to all individuals (solid lines) or to only seronegatives (dashed lines), inclusive of 96% serotest sensitivity, 99% specificity (*35*), and approximately three months of seroreversion (*32*) (see Methods). Shown: U.S. contact patterns and demographics (*22, 23*); all-or-nothing and transmission-blocking vaccine with 90% vaccine efficacy. See Figs. S12 and S13 for lower and higher seroprevalence examples, respectively.

## Discussion

This study demonstrated the use of an age-stratified modeling approach to evaluate and compare vaccine prioritization strategies for SARS-CoV-2. After accounting for country-specific age structure, age-contact structure, infection fatality rates, and seroprevalence, as well as the age-varying efficacy of a hypothetical vaccine, we found that across countries those aged 60 and older should be prioritized to minimize deaths, assuming a return to high contact rates and pre-pandemic behavior during or after vaccine rollout. This recommendation is robust because of the dramatic differences in IFR by age. Our model identified three general regimes in which prioritizing adults aged 20-49 would provide greater mortality benefits than prioritizing older adults. One such regime was in the presence of substantial transmission-mitigating interventions (*R*_0_ = 1.15) and a vaccine with 80% or higher transmission blocking effects. A second regime was characterized by substantial transmission-mitigating interventions (*R*_0_ = 1.15) and either rollout speeds of at most 0.2% per day or vaccine supplies of at most 25% of the population. The third regime was characterized by vaccines with very low efficacy in older adults, very high efficacy in younger adults, and declines in efficacy starting at age 59 or 69. The advantage of prioritizing all adults or adults 20-49 vs. adults 60+ was small under these conditions. Thus, we conclude that for mortality reduction, prioritization of older adults is a robust strategy that will be optimal or close to optimal to minimize mortality for virtually all plausible vaccine characteristics.

In contrast, the ranking of infection-minimizing strategies for mid-epidemic vaccination led to consistent recommendations to prioritize adults 20-49 across efficacy values and countries. For pre-transmission vaccination, prioritization shifted toward children and teenagers for leaky vaccine efficacies 50% and below, in line with prior work (*7*), as well as for vaccines with weak transmission-blocking properties. Because a vaccine is likely to have properties of both leaky and all-or-nothing models, empirical data on vaccine performance could help resolve this difference in model recommendations, although data are difficult to obtain in practice [see, e.g. (*36, 37*)].

It is not yet clear whether the first-generation of COVID-19 vaccines will be approved everywhere for the elderly or those under 18 (*38–40*). While our conclusions assumed that the vaccine would be approved for all age groups, the evaluation approaches introduced here can be tailored to evaluate a subset of approaches restricted to those within the age groups for which a vaccine is licensed, using open-source tools such as those that accompany this study (*21*). Furthermore, while we considered three possible goals of vaccination—minimizing cumulative incidence, mortality, or YLL—our framework can be adapted to consider goals such as minimizing hospitalizations, ICU occupancy (*7*) or economic costs (*10*).

We demonstrated that there is value in pairing individual-level serological tests with vaccination, even when accounting for the uncertainties in seroprevalence estimates (*34*) and seroreversion (*32*). The marginal gain in effective vaccine supply, relative to no serological testing, must be weighed against the challenges of serological testing prior to vaccination. Serostatus itself is an imperfect indicator of protection, and the relationship of prior infection, serostatus, and protection may change over time (*10, 29, 32, 33*). Delays in serological tests results would impair vaccine distribution, but partial seronegative-targeting effects might be realized if those with past PCR-confirmed infections voluntarily deprioritized their own vaccinations.

The best performing strategies depend on assumptions about the extent of a population’s interactions. We used pre-pandemic contact matrices (*22*), reflecting the goal of a return to pre-pandemic routines once a vaccine is available, but more recent estimates of age-stratified contact rates could be valuable in modeling mid-pandemic scenarios (*41, 42*). Whether pre-pandemic or mid-pandemic contact estimates are representative of contact patterns during vaccine rollout remains unknown and may vary based on numerous social, political, and other factors. The scenarios modeled here did not incorporate explicit non-pharmaceutical interventions, which might persist if vaccination coverage is incomplete, but are implicitly represented in Scenario 1 (*R*_0_ = 1.15).

Our study relies on estimates of other epidemiological parameters. In local contexts, these include age-structured seroprevalence and IFR, which vary by population (*19, 20, 43*). Globally, key parameters include the degree to which antibodies protect against reinfection or severity of disease and relative infectiousness by age. From vaccine trials, we also need evidence of efficacy in groups vulnerable to severe outcomes, including the elderly. Additionally, it will be critical to measure whether a vaccine that protects against symptomatic disease also blocks infection and transmission of SARS-CoV-2 (*44*).

The role of children during this pandemic has been unclear. Under our assumptions about susceptibility by age, children are not the major drivers of transmission in communities, consistent with emerging evidence (*12*). Thus, our results differ from the optimal distribution for influenza vaccines, which prioritize school-age children and adults age 30-39 (*5*). However, the relative susceptibility and infectiousness of SARS-CoV-2 by age remain uncertain. While it is unlikely that susceptibility to infection conditional on exposure is constant across age groups (*12*), we ran our model to test the sensitivity of this parameter. Under the scenario of constant susceptibility by age, vaccinating those under 20 has a greater impact on reducing cumulative cases than those 20-49 (Figs. S14, S15).

Our study is subject to a number of limitations. First, our evaluation strategy focuses on a single country at a time, rather than on between-population allocation (*45*). Second, we only consider variation in disease severity by age. However, other factors correlate with disease outcomes, such as treatment and healthcare access and comorbidities, which may correlate with factors like rural vs urban location, socioeconomic status, sex (*46, 47*), and race and ethnicity (*48*), that are not accounted for in this study. Inclusion of these factors in a model would be possible, but only with statistically sound measurements of both their stratified infection risk, contact rates, and disease outcomes. Even in the case of age stratification, contact surveys have typically not surveyed those 80 years and older, yet it is this population that suffers dramatically more severe COVID-19 disease and higher infection fatality rates. We extrapolated contact matrices to those older than 80, but direct measurements would be superior. Last, our study focused on guiding strategy rather than providing more detailed forecasting or estimates (*10*). As such, we have not made detailed parameter fits to time series of cases or deaths, but rather have used epidemiologic models to identify robust strategies across a range of transmission scenarios.

Our study also considers variation in disease risk only by age, via age-structured contact matrices and age-specific susceptibility, while many discussions around COVID-19 vaccine distribution have thus far focused on prioritizing healthcare or essential workers (*49, 50*). Contact rates, and thus infection potential, vary greatly not only by occupation and age but also by living arrangement (e.g., congregate settings, dormitories), neighborhood and mobility (*51–54*), and whether the population has a coordinated and fundamentally effective policy to control the virus. With a better understanding of population structure during the pandemic, and risk factors of COVID-19, these limitations could be addressed. Meanwhile, the robust findings in favor of prioritizing those age groups with the highest IFR to minimize mortality could potentially be extended to prioritize those with comorbidities that predispose them to a high IFR, since the strategy of prioritizing the older age groups depends on direct rather than indirect protection.

Vaccine prioritization is not solely a question of science but a question of ethics as well. Hallmarks of the COVID-19 pandemic, as with other global diseases, are inequalities and disparities. While these modeling efforts focus on age and minimizing incidence and death within a simply structured population, other considerations are crucial, from equity in allocation between countries to disparities in access to healthcare, including vaccination, that vary by neighborhood. Thus, the model’s simplistic representation of vulnerability (age) should be augmented by better information on the correlates of infection risk and severity. Fair vaccine prioritization should avoid further harming disadvantaged populations. We suggest that, after distribution, pairing serological testing with vaccination in the hardest hit populations is one possible equitable way to extend the benefits of vaccination in settings where vaccination might otherwise not be deemed cost-effective.

## Data Availability

Contact matrices, demographic data and model parameters are available via the cited references in the manuscript.

## Acknowledgements

The authors wish to thank Sereina Herzog, Mark Jit, Jacco Wallinga, and Helen Johnson for their feedback.

## Author Contributions

KMB, SMK, ML, SC, YHG and DBL conceived of the study. KMB and DBL performed the analyses. KMB and KR generated all figures. KR created interactive visualization tools. All authors wrote and revised the manuscript.

## Data and materials availability

Reproduction code is open source and provided by the authors (*21*).

## License

This work is licensed under a Creative Commons Attribution 4.0 International (CC BY 4.0) license, which permits unrestricted use, distribution, and reproduction in any medium, provided the original work is properly cited. To view a copy of this license, visit https://creativecommons.org/licenses/by/4.0/. This license does not apply to figures/photos/artwork or other content included in the article that is credited to a third party; obtain authorization from the rights holder before using such material.

## Supplementary Materials

### Materials and Methods

#### Susceptible Exposed Infectious Recovered (SEIR) Model Overview

We used a continuous-time ordinary differential equations (ODE) compartmental model stratified by age. Across all model variations and analyses, we simulated 365 days of dynamics, corresponding to the first-year phase of vaccine prioritization. All individuals were assumed to be initially susceptible, unless they had been effectively vaccinated or had naturally acquired immunity, which was considered to be protective in this model. Susceptible people (*S*) transition to the exposed state (*E*) after contact with an infectious individual. After a latent period, exposed individuals become infectious (*I*). After an infectious period, individuals move to a recovered state (*R*) or die. We assume that recovered individuals are no longer infectious and are immune to reinfection over the duration of simulations (up to 365 days). The duration of time spent in compartments *E* and *I*, in expectation, are specified in Table S1. Model equations were solved using *lsoda* ODE solver from the package *deSolve*, R version 3.6.0 (*55*). Fig. S1 shows model schematic diagrams for the variations of the SEIR model considered in this manuscript. The force of infection, *λ*_*i*_ for a susceptible individual in age group *i* is

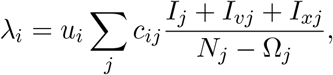

where *u*_*i*_ is the probability of a successful transmission given contact with an infectious individual and *c*_*ij*_ is the number of age-*j* individuals that an age-*i* individual contacts per day. The term (*I*_*j*_ + *I*_*vj*_ + *I*_*xj*_)*/*(*N*_*j*_ Ω_*j*_) is the probability that a random age-*j* individual is infectious, where *I*_*j*_ is the number of individuals who are unvaccinated and infectious, *I*_*vj*_ is the number of individuals who are vaccinated yet infectious, *I*_*xj*_ is the number of individuals who are ineligible for vaccination (e.g. due to hesitancy or due to a positive serological test) and infectious, *N*_*j*_ is the total number of individuals in group *j* and Ω_*j*_ is the number of individuals from group *j* who have died. To calculate the basic reproductive number, *R*_0_, we define the next-generation matrix as

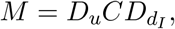

where *D*_*u*_ is a diagonal matrix with diagonal entries *u*_*i*_, *C* is the country-specific contact matrix, and *D*_*d*__*I*_ is a diagonal matrix with diagonal entries *d*_*I*_, the infectious period. *R*_0_ is the absolute value of the dominant eigenvalue of *M*. Age-stratified susceptibility values were drawn from literature estimates (*13*). Table S1 details all parameters used in this manuscript and their sources.

### Incorporation of Vaccine Hesitancy

To incorporate vaccine hesitancy, we limited vaccine uptake such that at most 70% of any age group was eligible to be vaccinated (*24*). To implement this restriction, 30% of each compartment for each age group was initialized as ineligible for vaccination. These individuals were tracked using compartments *S*_*x*_, *E*_*x*_, *I*_*x*_, and *R*_*x*_ (Fig. S1). Initial conditions, inclusive of 30% vaccine hesitancy, are listed in Table S2.

### Incorporation of Vaccination, Vaccine Rollout, and Vaccine Efficacy

In the simplest version of the model, the vaccine is assumed to be transmission- and infection-blocking, and to work with variable efficacy. We considered two classes of scenarios. In the first class of scenarios, vaccinations are given in advance of model dynamics, which we call pre-transmission vaccination. In the second class of scenarios, vaccinations are rolled out at the same time as the model dynamics, which we call continuous rollout vaccination.

In continuous rollout scenarios (Scenarios 1 and 2), vaccine rollout was parameterized by the percentage of the total population that was vaccinated in each day of simulation, with values ranging from 0.05% to 1% of total population (see Fig. S6). Scenarios 1 and 2 of the Main Text consider rollout speeds of 0.2% per day. The prescribed number of individuals received the effects of vaccination in simulations prior to the start of each day, such that disease dynamics proceeded in continuous time while vaccine rollout was computed in discrete steps. We did not include an explicit delay between vaccination and protection, and therefore our approach may be thought of as either a model for a vaccine with immediate protective effects, or as a model in which the time of protection is explicitly modeled and injections are thus implicitly assumed to precede said protection. In continuous rollout scenarios, the model was seeded with 0.25% of individuals in each age group exposed and 0.25% of individuals in each age group infectious.

In pre-transmission vaccination scenarios (Scenario 3 and 4), all the available vaccines were distributed at the initial time step, prior to the epidemic. To incorporate vaccinations, we initialized the model by dividing the total population of each age group between the susceptible compartment (*S*) and vaccinated compartment (*V* or *S*_*v*_), according to the vaccine prioritization strategy and number of vaccines available. In pre-transmission vaccination scenarios, the model was seeded with one infectious person in *I* and one infectious person in *I*_*x*_ in each age group. Scenarios 3 and 4 of the Supplementary Text use pre-transmission rollout.

We considered two ways to implement vaccine efficacy (*ve*): as an all-or-nothing vaccine, where the vaccine provides perfect protection to a fraction *ve* of individuals who receive it, or as a leaky vaccine, where all vaccinated individuals have reduced probability *ve* of infection after vaccination (see Supplementary Text). We ran simulations with both types of vaccine efficacy; Figures in the Main Text show results only for all-or-nothing vaccines.

To incorporate age-dependent vaccine efficacy, we parameterized the relationship between age and vaccine efficacy via an age-efficacy curve with (i) a baseline efficacy, an age at which efficacy begins to decrease (hinge age), and a minimum vaccine efficacy *ve*_*m*_ for adults 80+ (Fig. 3A). We assumed that *ve* is equal to the baseline value for all ages younger than the hinge age, then decreases stepwise in equal increments for each decade to the specified minimum *ve*_*m*_ for the 80+ age group. To determine whether there exists a *ve*_*m*_ such that the mortality-minimizing strategy switches from directly vaccinating adults 60+ to an alternative strategy, we used the bisection method (*56*).

### Incorporation of Existing Seroprevalence

To incorporate existing seroprevalence estimates and compare areas with differing naturally-acquired immunity, we used data and seroprevalence estimates from Connecticut [low seroprevalence; (*31*)] and New York City [moderate seroprevalence; (*30*)]. To model high seroprevalence, we simulated an unmitigated epidemic with *R*_0_ = 2.6 until 40% cumulative incidence was reached. Seroprevalence was implemented by moving the proportion of seropositive individuals from each age group into the recovered compartment prior to forward simulations that incorporated vaccination.

The model’s implementation of vaccination depended on whether the vaccine was rolled out during on-going transmission or prior to transmission. For pre-transmission vaccination without consideration of serostatus, *v*_*i*_ doses were given to each population group *i*, a fraction *θ*_*i*_ of whom were already recovered. Thus, the total number of individuals eligible for vaccination were *S* + *R*, assuming currentely infected individuals do not seek vaccination. In scenarios where vaccination was targeted only at seronegative individuals, simulations were conducted with sensitivity 70% and specificity 99%, incorporating the performance of a hypothetical Euroimmun IgG test (*35*) and a 25% reduction in sensitivity due to seroreversion (*32*). Details of continuous vaccine rollout with dose redirection using an imperfect serological test can be found in Supplementary Text.

### Calibration to achieve target R_0_

Models were calibrated to achieve the target *R*_0_ by multiplying the next-generation matrix by a constant to achieve the desired dominant eigenvalue, i.e. *R*_0_. Because the constant factors out of the next-generation matrix equation, this may be mathematically interpreted as scaling up or down either the contact rates *C* or susceptibilities *u*. All model calibration was performed prior to the inclusion of vaccination, meaning that the reproductive number *R* in the first days of a simulation may differ from *R*_0_ depending on the scenario considered. Values of *R*_0_ studied ranged from 1.1 to 2.6. When incorporating seroprevalence, calibration was performed after the inclusion of seroprevalence.

### Measurement of outcomes: infections, deaths, and years of life lost

We ran simulations for 365 days from the date of the first vaccination to focus on the early prioritization phase of the COVID-19 vaccination programs. To compare the impact of different vaccination prioritization strategies, we calculated the cumulative number of infections and deaths. Infected individuals either move to the recovered or dead compartment, according to the age-dependent IFR (*19*) (see Fig. S1). The cumulative number of new infections since the onset of vaccine rollout is the total number of recovered and dead at the end of the simulation minus the initial number of seropositive individuals, Σ*_i_ R*_*i*_ + *R*_*x,i*_ + *R*_*v,i*_ + Ω_*i*_ *θ*_*i*_*N*_*i*_. The total number of estimated deaths was the number of people in the dead compartment at the end of the simulation. To calculate years of life lost (YLL) due to a death at a particular age, we multiplied standard life expectancy (SLE) by the number of deaths per age bin. We used the country-specific SLE estimates from the WHO Global Health Observatory (*57*), aggregated into age bins by decade using 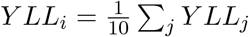 where *j* are the ages corresponding to decadal age bin *i*.

### Contact Matrices and Demographics

Country-specific contact matrices include four types of contact: home, work, school, and other (*22*). In all simulations, we used total contact matrices, equivalent to the sums of the four contact types. Age demographics in all simulations were taken from the UN World Population Prospects 2019 for each country (*23*). Age bins in each case were originally provided in 5-year increments, which were then combined into 10-year increments by addition. For instance, the number of individuals between 20 and 29 was the sum of individuals 20-24 and 25-29. The number of individuals 80 years and older was calculated as the sum of all age bins greater than 80.

We made two adaptations to existing contact matrices (*22*). First, we combined their five year age bins into ten year bins. Each entry *x*_*ij*_ in the original matrices corresponds to the number of individuals of age-group *j* that a person in age group *i* typically comes into contact with. Thus, for a country with population fraction *d*_*i*_ in age group *i*, the combined contact matrix entries are given by

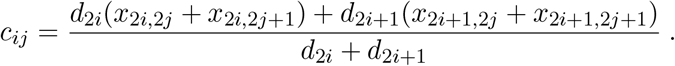

Second, we extrapolated matrices to include individuals 80+. To extrapolate, we copied the contact rates from 70-79 y to our new row and column for 80+, along the diagonal. Then we filled in the end of our new row and column with the 70-79 y contact rates with 0-9 y, assuming interactions with 0-9 y are similar for people 70+. Lastly, to account for increased housing in long term living facilities for 80+ y, we decreased their contacts for 0-60 y by 10% and added it to the 70 and 80 y contacts. Thus, 80+ year-olds have the same total number of contacts as 70-79 year-olds, but relatively fewer among 0-69-year-olds and proportionally more among 70+ year-olds.

**Figure S1:**
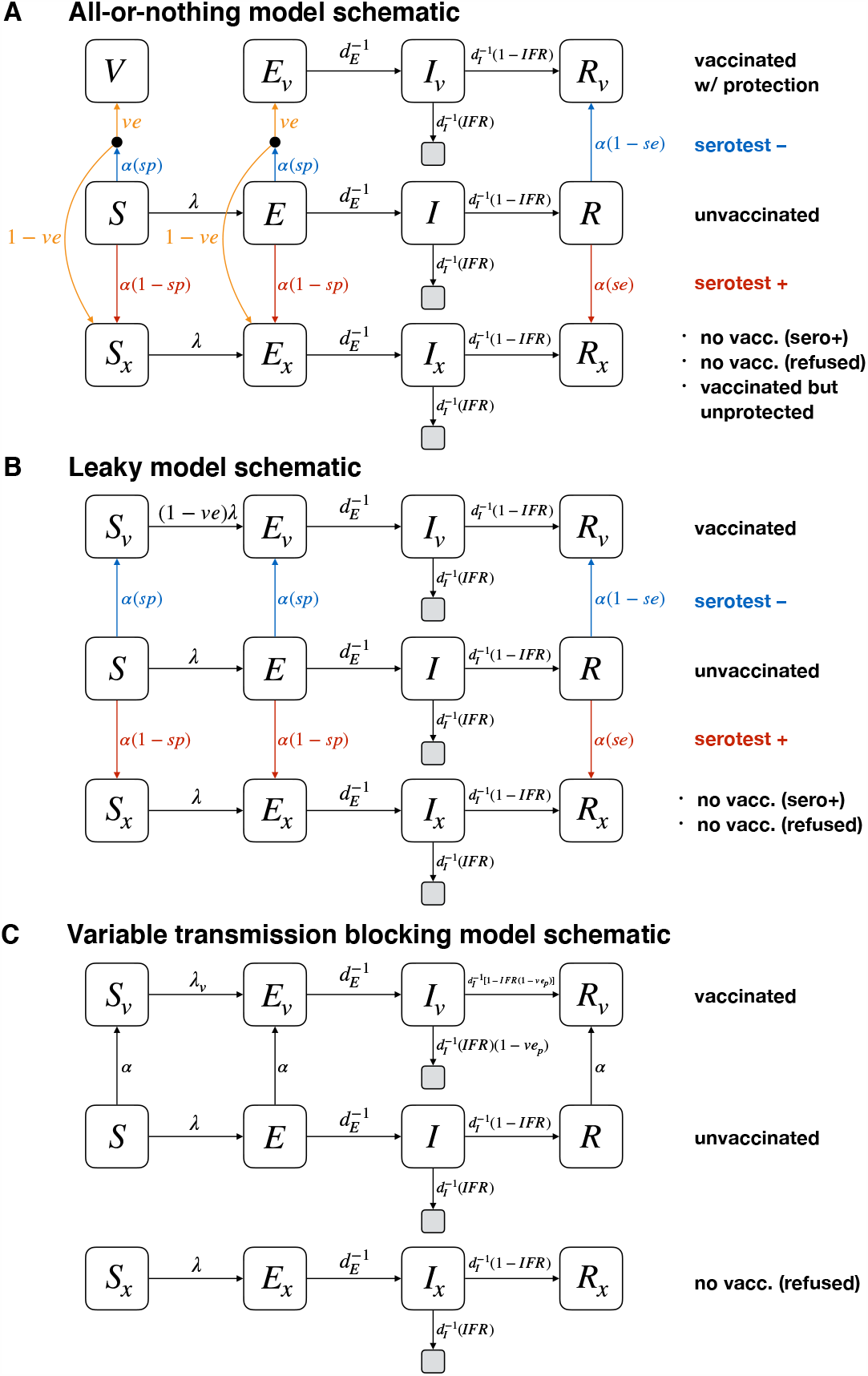
Schematics for vaccine modeling framework. Diagrams show compartmental models and transition rates for the (A) all-or-nothing, (B) leaky, and (C) variable transmission-blocking vaccine models used in this manuscript. *S, E, I*, and *R* represent susceptible, exposed, infectious, and recovered compartments; *V* represents a perfectly protected and vaccinated compartment; subscripts of *v* and *x* denote those who have been vaccinated with protection (*v*), and those who will either not be vaccinated (vaccine refusal or positive serotest) or have been vaccinated but without protection (*x*). Grey unlabeled compartments represent death. The incorporation of a point-of-care serological test can be included by using known sensitivity *se* and specificity *sp*, or can be excluded by setting *se* = 0 and *sp* = 1 (a convenient mathematical representation of the no-test scenario is simply a test that always returns a negative result). Vaccine rollout count *α* is given by *α* = *n*_vax_*/* [(*S* + *E*)*sp* + *R*(1 − *se*)] where *n*_vax_ is the total number of vaccines to be rolled out in a particular day. All compartments are stratified by age, in the text, with index *i*. See text for details and initial conditions.

## Supplementary Text

### SEIR Model Modifications

The flexible age-stratified SEIR model framework allowed us to model (i) leaky or all-or-nothing vaccine efficacy, (ii) variable rollout speeds, (iii) point-of-care reprioritization of vaccines using an imperfect serological test, and (iv) the possibility of only partial transmission blocking effects, through straight-forward modifications. The framework is shown in Fig. S1, and explicitly tracks individuals who were vaccinated and protected, vaccinated but not protected (all-or-nothing model only), considered for vaccination but did not receive a dose due to a positive serological test, and those who were not vaccinated or considered for vaccination. These four modes of operation are described in the subsections below.

### Implementing vaccine efficacy: leaky vs all-or-nothing

A vaccine with imperfect efficacy can be modeled as either a “leaky” vaccine, where all vaccinated individuals are *ve* protected against infection (Fig. S1A), or an “all-or-nothing” vaccine, where a fraction *ve* of vaccinated individuals are perfectly protected while the remaining 1 *-ve* individuals gain no protection (Fig. S1B). We considered both model implementations of vaccine efficacy, showing results for the all-or-nothing model in the Main Text and the leaky model in Supplementary Figures, as indicated in figure captions.

### Implementing variable vaccine rollout rate

We allowed vaccines to be distributed (or “rolled out”) at different rates by parameterizing the number of vaccines available in each simulated day, *n*_vax_. In each simulated day, exactly *n*_vax_ doses are distributed, according to the prioritization strategy, prior to calculating new exposures, infections, and recoveries. As a result, pre-transmission vaccination, in which all doses are used prior to SEIR dynamics, can be implemented by setting *n*_vax_ equal to the total number of doses available. In all scenarios, when all doses have been used, *n*_vax_ = 0 for all timesteps thereafter.

### Implementing point-of-care dose redirection with imperfect serological tests

The modeling framework depicted in Fig. S1 allows for the point-of-care reprioritization of tests by using the outcome of an imperfect serological test with sensitivity *se* and specificity *sp*. We assume that only those who test negative receive a vaccine, but that this population may consist of true negatives from the *S* or *E* compartments or false negatives from the *R* compartment. By tracking all such individuals, we prevent the model from vaccinating the same person twice. Similarly, we assume that those who test positive do not receive a vaccine at any point after testing positive. These include false positives from the *S* and *E* compartments and true positives from the *R* compartment.

The numbers of individuals who receive a vaccination each timestep, given a supply of *n*_vax_ to be allocated in that timestep, are

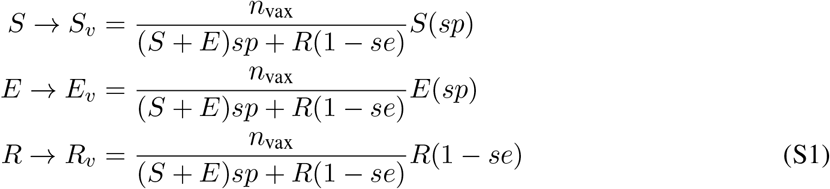

while the numbers of individuals who are tested but are excluded from vaccination are

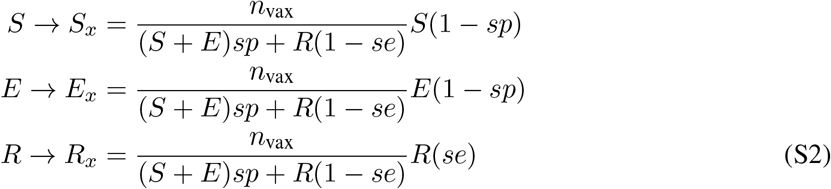

How many more susceptible individuals get vaccinated when point-of-care reprioritization is used? This can be computed *a priori* for pre-transmission rollouts in which a fraction *θ*_*i*_ of individuals in subpopulation *i* are truly seropositive and thus treated as recovered. If we further assume no ongoing transmission, then *S* = (1 − *θ*_*i*_)*N* (1 − *vh*) where *vh* is the fraction of each age group that is vaccine hesitant, and the number of susceptible individuals vaccinated is

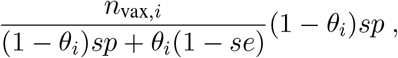

while the number of susceptible individuals vaccinated without point-of-care serology is

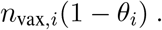

We note that vaccination without a serotest is equivalent to setting *se* = 0 and *sp* = 1, i.e. considering a “test” that always returns a negative result. The relative increase in the number of individuals vaccinated is thus

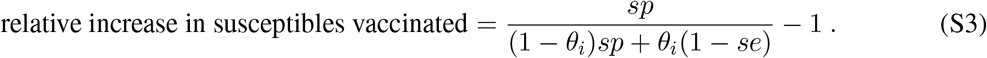

Recent work suggests that seroprevalence decreases over time due to seroreversion (*32*). As a consequence, Eq. S3 and the calculations used to derive it must be interpreted with care. If *θ*_*i*_ represents the current seroprevalence, Eq. (S3) may be used with the published sensitivity and specificity of a serological test (e.g. 99% specificity and 96% sensitivity for the Euroimmun IgG (*35*)). However, if *θ*_*i*_ represents a past measurement of seroprevalence or an estimate of cumulative incidence, then the sensitivity of a test to identify individuals with past exposure will be diminished, with estimates suggesting seroreversion of 27% of individuals over 3 months (*32*). To more conservatively estimate the impact of using serology for dose redirection, a test with 70% sensitivity and 99% specificity in a population with a past measurement of 25% seropositivity would lead to a 21.1% increase in susceptibles vaccinated. When vaccine rollout is not pre-transmission, and instead continues alongside transmission, *a priori* calculations of this type are not possible.

### Implementing vaccines with imperfect transmission blocking

We considered a vaccine that prevents severe manifestations of COVID-19 infection, including death, with 90% efficacy, but imperfectly blocks transmission of SARS-CoV-2 (Fig. S1C). To model such a vaccine, we modify the leaky vaccine model by introducing three different mechanisms for vaccine efficacy: *ve*_*S*_ is the efficacy of the vaccine to decrease susceptibility; *ve*_*I*_ is the efficacy of the vaccine to decrease infectiousness; and *ve*_*P*_ is the efficacy of the vaccine to decrease the likelihood that the infection progresses to severe disease and death.

In this model, the infectiousness of vaccinated individuals is decreased by a factor of 1 *-ve*_*I*_. Second, the susceptibility of vaccinated individuals is decrease by a factor of 1 *-ve*_*S*_. This results in a force of infection for unvaccinated individuals of

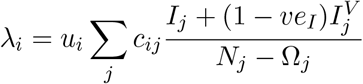

and a force of infection for vaccinated individuals of

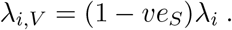

These values of *λ*_*i*_ replace previous values of *λ*_*i*_ for *S* → *I* transitions, and values of *λ*_*i,V*_ replace values of (1 − *ve*)*λ*_*i*_ for *S*_*V*_ → *I*_*V*_ transitions (see Fig. S1). Finally, the fatality rate, conditioned on infection, is multiplied by a factor of 1 − *ve*_*P*_.

The impact of this variable-transmission blocking model on minimizing incidence and mortality are shown in Supplementary Fig. S10 for *ve*_*S*_ = 0, *ve*_*P*_ = 90% and *ve*_*I*_ = 0 *-*100%. Note that when *ve*_*S*_ = *ve*_*I*_ = 0, there are no indirect effects of vaccination. Finally, we note that when considering a leaky vaccine with different effects on infection, transmission, and progression, the parameterization could be accomplished in any of several ways. Ours makes the choice for simplicity to consider the vaccine’s entire effect to be on infectiousness and progression.

### Results for Pre-Transmission Vaccine Rollout

The governments and residents of New Zealand, Taiwan, and South Korea, among others, have been dramatically more successful at mitigating the spread of SARS-CoV-2 than the rest of the world. As a result, widespread vaccination is likely to occur prior to the reopening of external borders and the relaxation of restrictions and testing of incoming travellers. To better model these particular scenarios, we conducted additional analyses using pre-pandemic contact matrices but implemented vaccination prior to the start of model dynamics. Here, we review our findings for such pre-transmission vaccination with *R*_0_ = 1.5 (Scenario 3) and *R*_0_ = 2.6 (Scenario 4).

Of the five strategies, direct vaccination of adults over 60 years (60+) nearly always reduced mortality and YLL more than the alternative strategies when transmission was high, across vaccine efficacies and supplies (*R*_0_ = 2.6; Scenario 4. Fig. S5). Exceptions occurred only vaccine efficacy was 80% or higher and supply was sufficient to cover over 45% of the population (Fig. S5). However, when transmission was lower (*R*_0_ = 1.5; Scenario 3), prioritizing adults 20-49 most reduced mortality in a similarly broad range of vaccine efficacies and supplies (Fig. S5).

Across countries, a general pattern emerged in which prioritization of adults 60+ was more often the mortality-minimizing strategy when transmission was higher and adults 20-49 when transmission was lower (Scenario 4 vs Scenario 3). However, for many vaccine supply levels and 90% vaccine efficacy, the prioritization of adults 20+ was also optimal (Fig. S9).

Across all scenarios, vaccine supplies, efficacies, and values of *R*_0_, the prioritization of adults 20-49 most reduced cumulative incidence in pre-transmission vaccination, with two exceptions. First, for high transmission (*R*_0_ = 2.6; Scenario 4), vaccination of those under 20 was most effective when efficacy was under 60%, a result restricted to the leaky vaccine model (Fig. S7). Second, for high transmission (*R*_0_ = 2.6; Scenario 4), vaccination of those under 20 was most effective when transmission-blocking efficacy was imperfect and vaccine supplies were low or moderate (Fig. S10).

Direct comparison of Scenario 2 (*R*_0_ = 1.5, rollout 0.2% per day) and Scenario 3 (*R*_0_ = 1.5, pre-transmission vaccination) shows that the ability to vaccinate prior to transmission markedly changes optimal strategies. These results were unaffected by modeling the vaccine as all-or-nothing (Fig. S5) or leaky (Fig. S7), except that for vaccine efficacy below 60% and high transmission (Scenario 4), prioritization of those under 20 most reduced transmission.

In general, pre-transmission vaccination markedly reduced both mortality and transmission, and expanded the vaccine supply and efficacy combinations for which prioritizing adults 20-49 most reduced mortality and YLL across all investigated values of *R*_0_ (Fig. S8), and across countries (Fig. S9), relative to mid-epidemic vaccine rollout.

**Figure S2:**
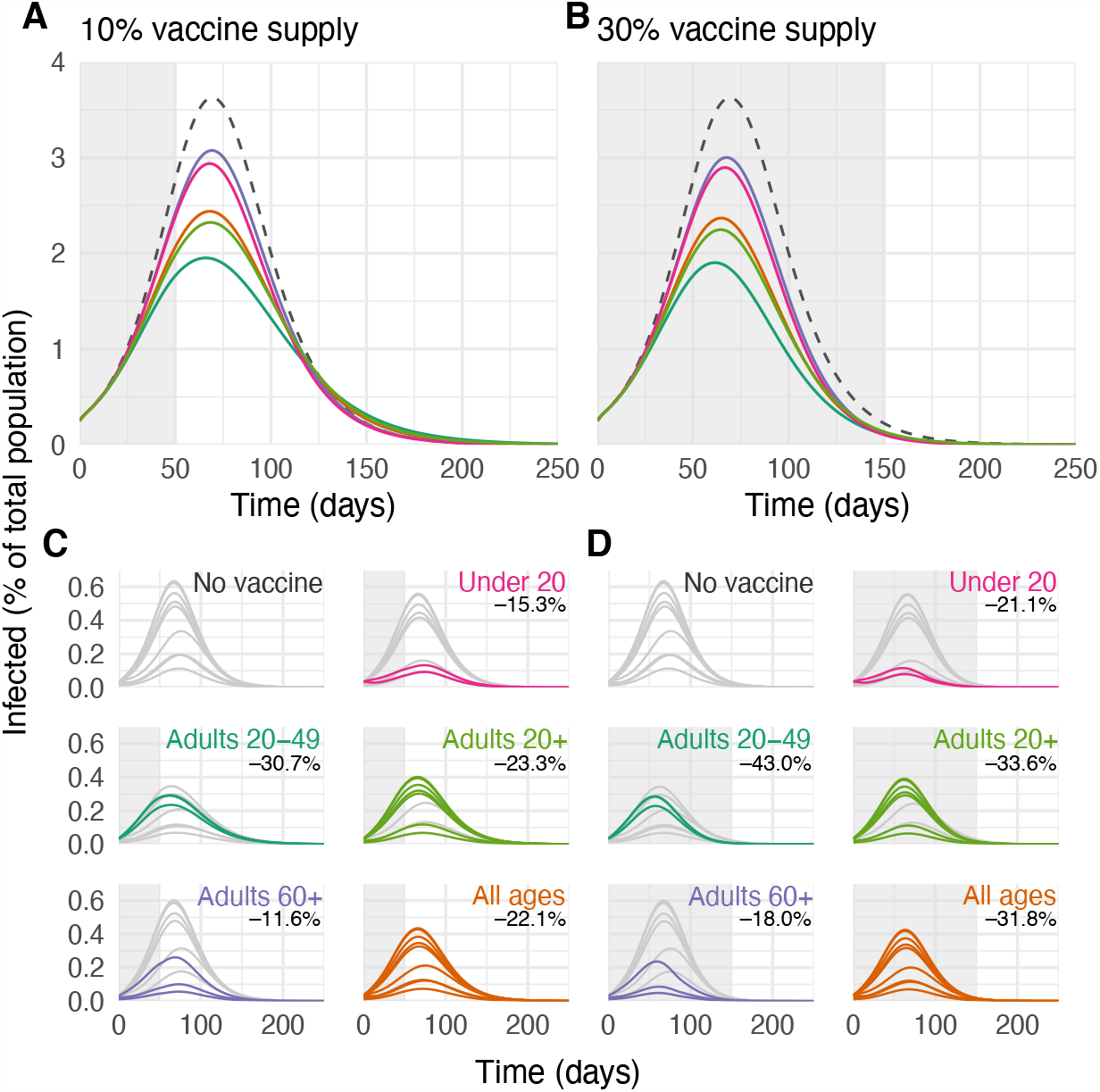
Example infection curves under varying vaccine supply and prioritization strategy. (A, B) The percentage of the total population infected over time is shown during simulations with no vaccines (grey dashed lines) and for five different prioritization strategies, with 10% (A) and 30% (B) vaccine supply. (C, D) The percentage of the total population infected over time, stratified into the decadal age groups used in simulations, are shown for both prioritized age groups (colored lines) and unvaccinated age groups (grey lines), with 10% (C) and 30% (D) vaccine supply. Annotations indicate the age groups targeted by each strategy and record the total reductions in infections after 365 simulated days. Shown: contact patterns and demographics of the United States (*22,23*); all-or-nothing and transmission blocking vaccine, *R*_0_ = 1.5, *ve* = 90%, rollout speed 0.2% of the population per day. See Figs. S3 and S4 for cumulative incidence and mortality curves.

**Figure S3:**
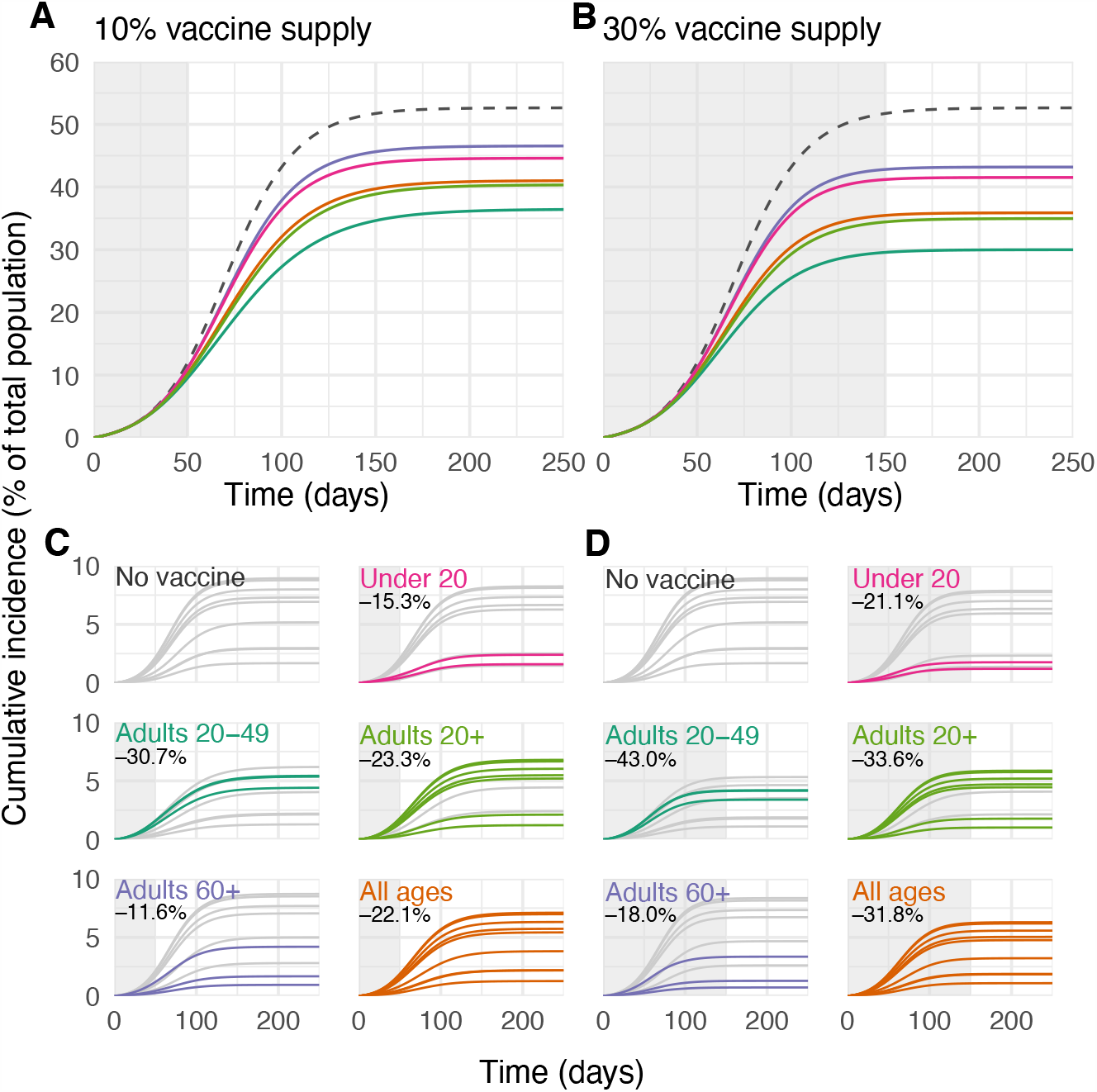
Example cumulative incidence curves under varying vaccine supply and prioritization strategy. (A, B) The percentage of the total number of infections over time is shown during simulations with no vaccines (grey dashed lines) and for five different prioritization strategies, with 10% (A) and 30% (B) vaccine supply. (C, D) The percentage of the total number of infections over time, stratified into the decadal age groups used in simulations, are shown for both prioritized age groups (colored lines) and unvaccinated age groups (grey lines), with 10% (C) and 30% (D) vaccine supply. Annotations indicate the age groups targeted by each strategy and record the total reductions in infections after 365 simulated days. Shown: contact patterns and demographics of the United States (*22, 23*); all-or-nothing and transmission blocking vaccine, *R*_0_ = 1.5, vaccine efficacy=90%, rollout speed 0.2% of the population per day. See Figs. S2 and S4 for infections and cumulative mortality curves.

**Figure S4:**
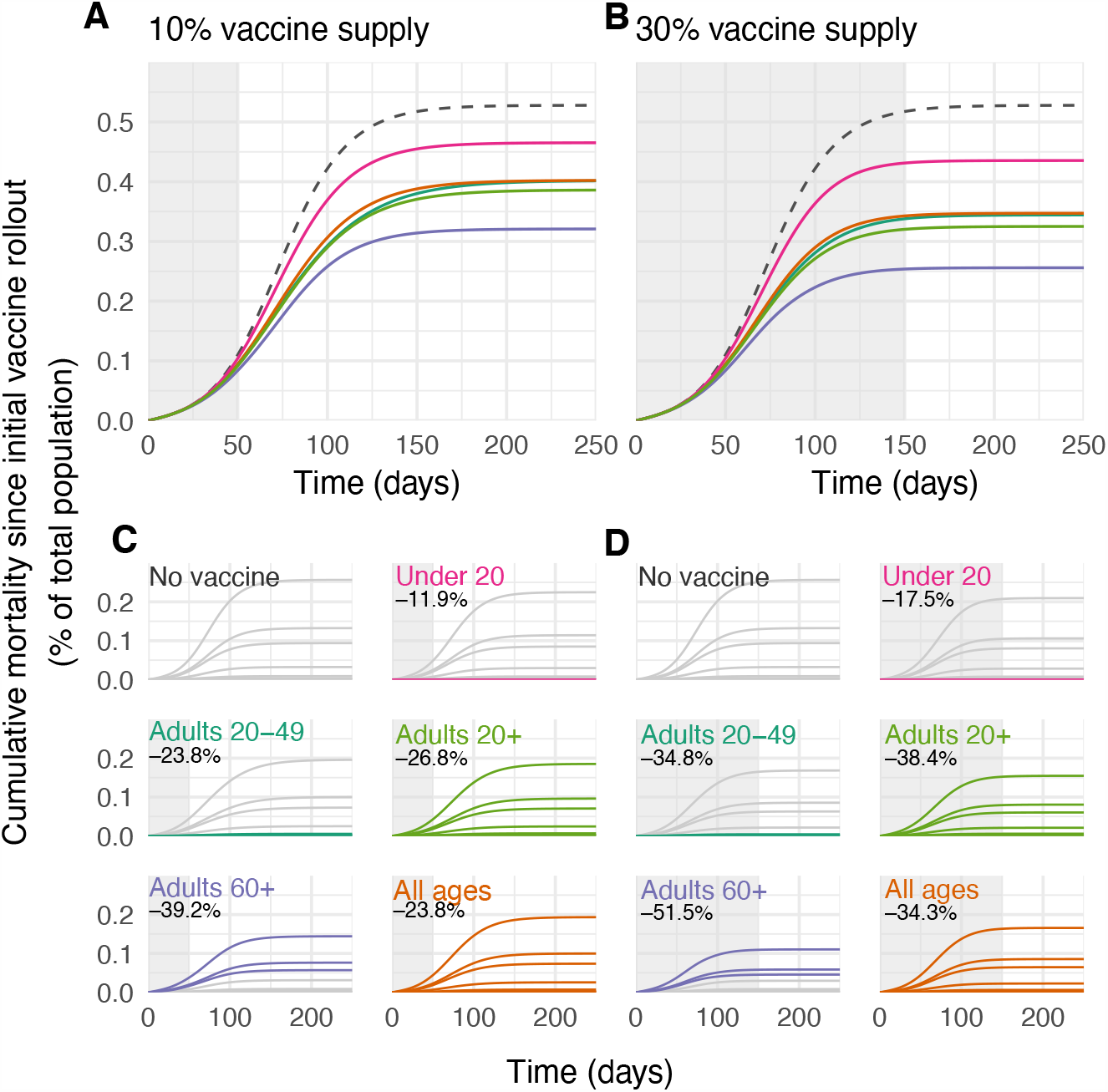
Example cumulative mortality curves under varying vaccine supply and prioritization strategy. (A, B) The cumulative mortality as a percentage of the total population is shown during simulations with no vaccines (grey dashed lines) and for five different prioritization strategies, with 10% (A) and 30% (B) vaccine supply. (C, D) The cumulative mortality as a percentage of the total population, stratified into the decadal age groups used in simulations, are shown for both prioritized age groups (colored lines) and unvaccinated age groups (grey lines), with 10% (C) and 30% (D) vaccine supply. Annotations indicate the age groups targeted by each strategy and record the total reductions in infections after 365 simulated days. Shown: contact patterns and demographics of the United States (*22, 23*); all-or-nothing and transmission blocking vaccine, *R*_0_ = 1.5, vaccine efficacy=90%, rollout speed 0.2% of the population per day. See Fig. S2 and S3 for infections and cumulative incidence curves.

**Figure S5:**
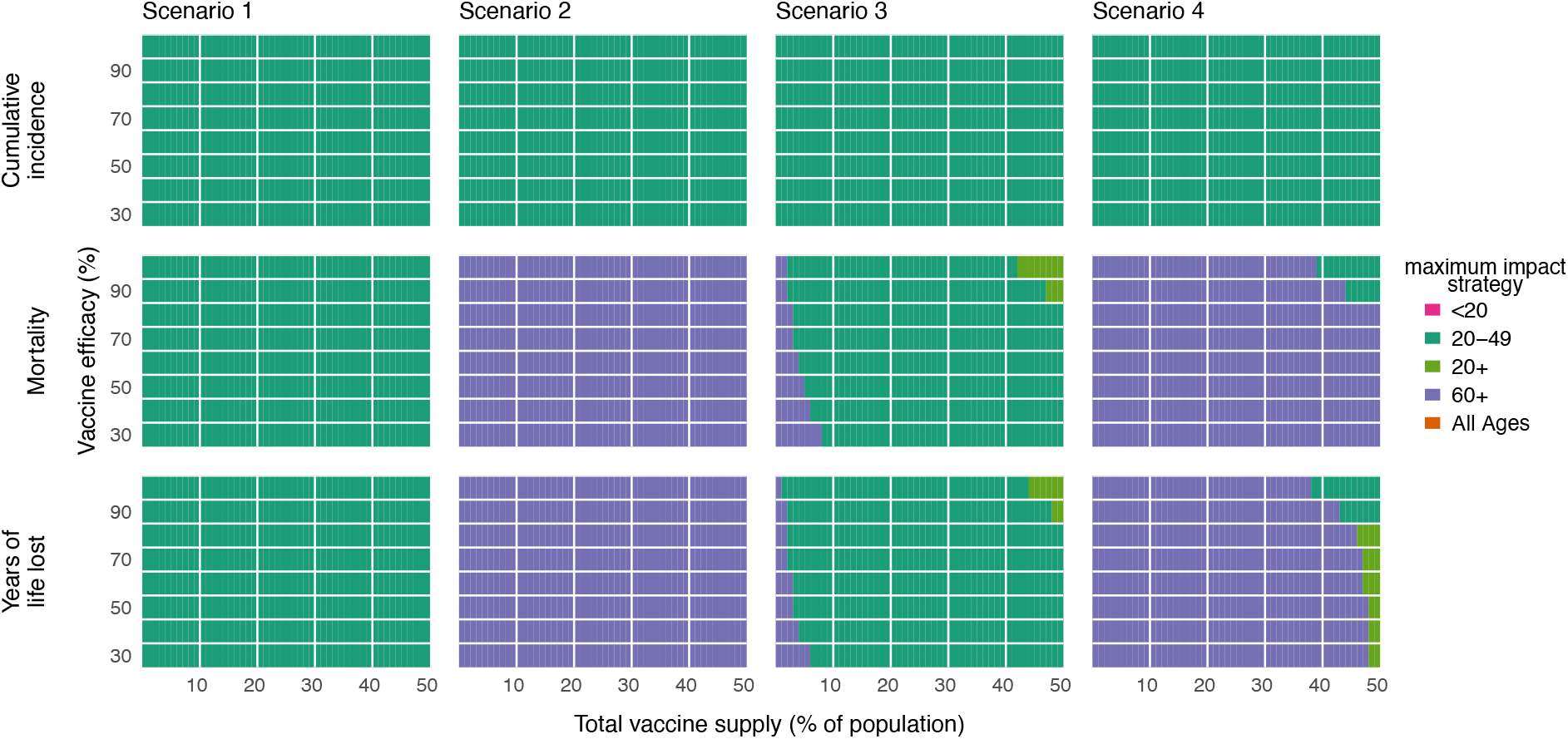
Impact of vaccine efficacy on maximum impact strategies (all-or-nothing vaccine). Heatmaps show the prioritization strategies resulting in maximum reduction of infections (top row), mortality (middle row), and years of life lost (bottom row) across Scenario 1 (0.2% rollout/day, *R*_0_ = 1.15; left column), Scenario 2 (0.2% rollout/day, *R*_0_ = 1.5; left-middle column), Scenario 3 (pre-transmission vaccination, *R*_0_ = 1.5; right-middle column), and Scenario 4 (pre-transmission vaccination, *R*_0_ = 2.6; right column). Each heatmap shows results from simulations varying vaccine supply and vaccine efficacy as indicated. Shown: contact patterns and demographics of the United States (*22, 23*); all-or nothing and transmission blocking vaccine. See Fig. S7 for leaky vaccine results.

**Figure S6:**
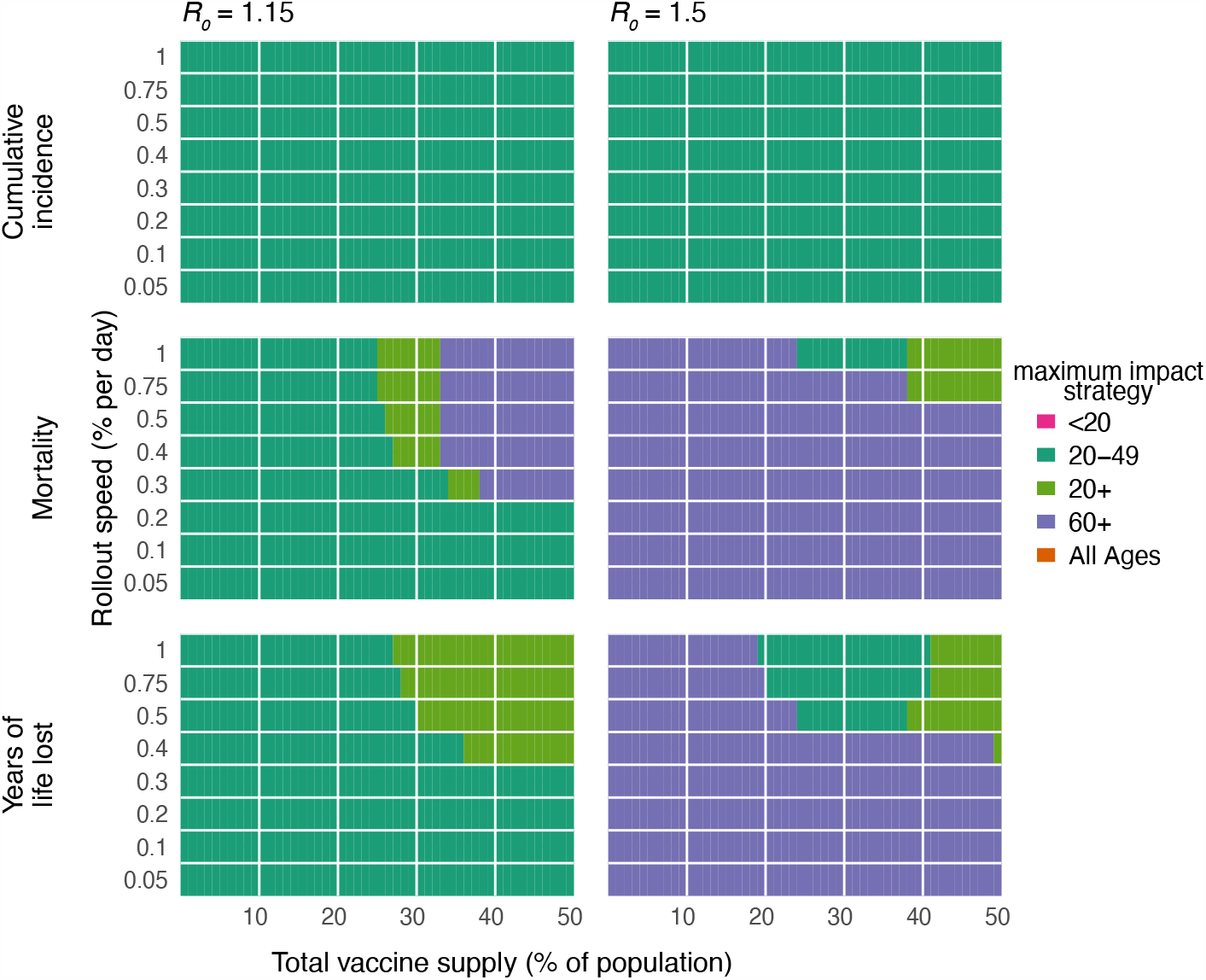
Impact of rollout speed on maximum impact strategies. Heatmaps show the prioritization strategies resulting in maximum reduction of infections (top row), mortality (middle row), and years of life lost (bottom row) for *R*_0_ = 1.15 (left column) or *R*_0_ = 1.5 (right column). Each heatmap shows results from simulations varying vaccine supply and the rollout speed of the vaccine, measured in the percentage of the total population vaccinated per day. Shown: contact patterns and demographics of the United States (*22, 23*); all-or nothing and transmission blocking vaccine, with vaccine efficacy = 90%.

**Figure S7:**
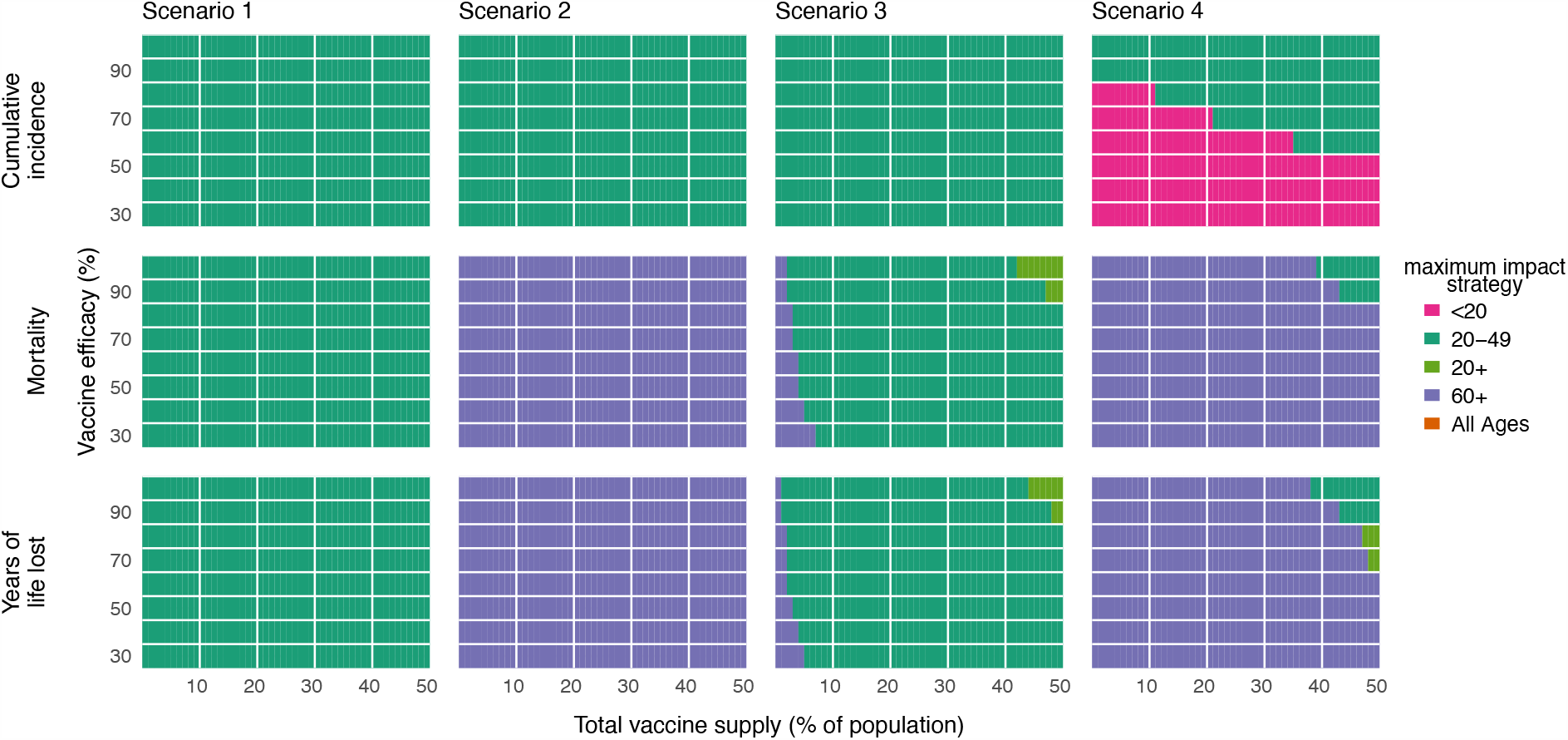
Impact of vaccine efficacy on maximum impact strategies (leaky vaccine). Heatmaps show the prioritization strategies resulting in maximum reduction of infections (top row), mortality (middle row), and years of life lost (bottom row) across Scenario 1 (0.2% rollout/day, *R*_0_ = 1.15; left column), Scenario 2 (0.2% rollout/day, *R*_0_ = 1.5; left-middle column), Scenario 3 (pre-transmission vaccination, *R*_0_ = 1.5; right-middle column), and Scenario 4 (pre-transmission vaccination, *R*_0_ = 2.6; right column). Each heatmap shows results from simulations varying vaccine supply and vaccine efficacy as indicated. Shown: contact patterns and demographics of the United States (*22, 23*); leaky and transmission blocking vaccine. See Fig. S5 for all-or-nothing vaccine results.

**Figure S8:**
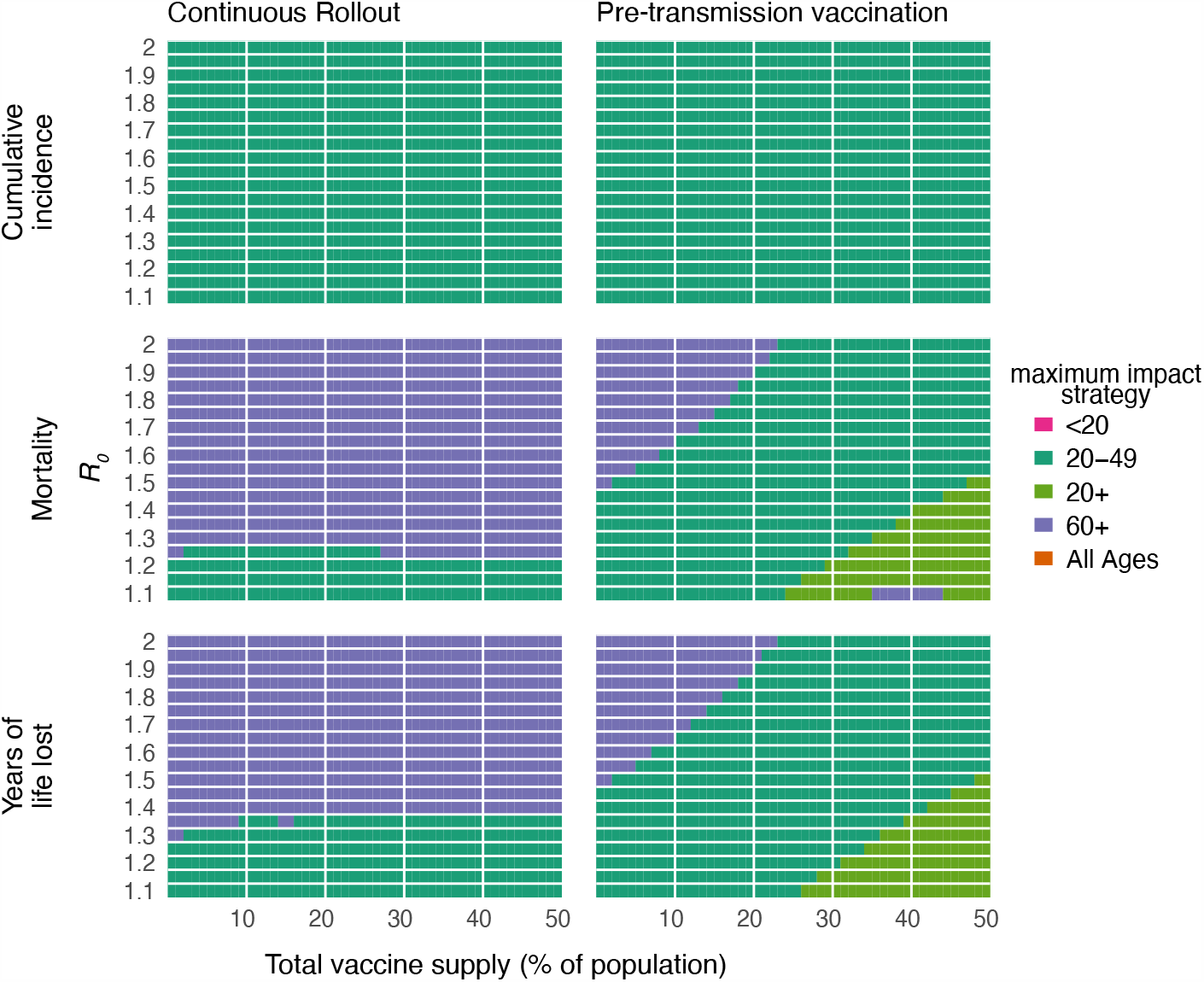
Impact of reproductive number R_0_ on maximum impact strategies. Heatmaps show the prioritization strategies resulting in maximum reduction of infections (top row), mortality (middle row), and years of life lost (bottom row) for a continuous rollout scenario (0.2% rollout/day; left column) or a pre-transmission rollout scenario (all doses administered prior to simulations; right column). Each heatmap shows results from simulations varying vaccine supply and the basic reproductive number *R*_0_. Shown: contact patterns and demographics of the United States (*22, 23*); all-or nothing and transmission blocking vaccine, with vaccine efficacy=90%.

**Figure S9:**
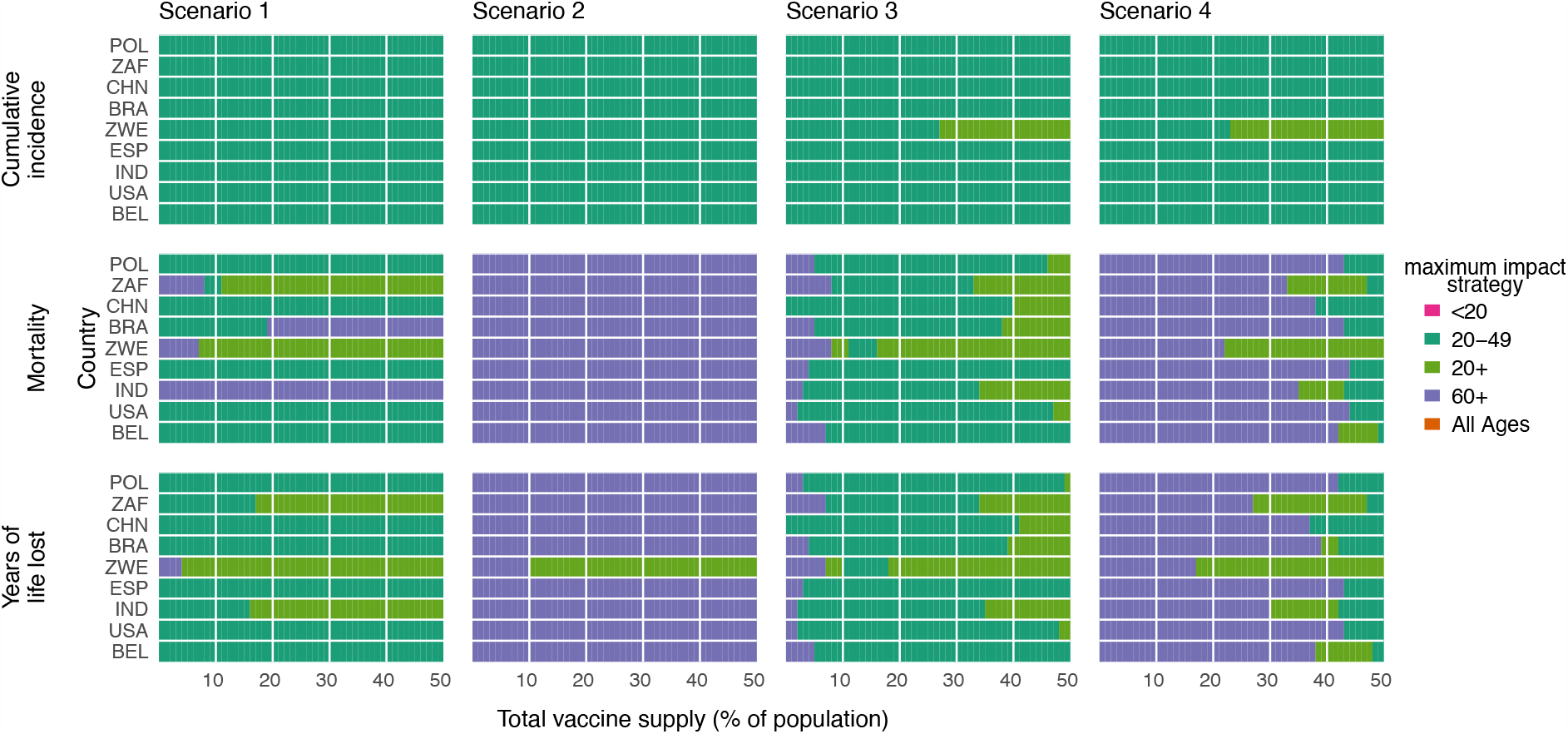
Impact of country demography and contact patterns on maximum impact strategies. Heatmaps show the prioritization strategies resulting in maximum reduction of infections (top row), mortality (middle row), and years of life lost (bottom row) across Scenario 1 (0.2% rollout/day, *R*_0_ = 1.15; left column), Scenario 2 (0.2% rollout/day, *R*_0_ = 1.5; left-middle column), and Scenario 3 (pre-transmission vaccination, *R*_0_ = 1.5; right-middle column), and Scenario 4 (pre-transmission vaccination, *R*_0_ = 2.6; right column). Each heatmap shows results from simulations varying vaccine supply and the country whose demographics and contact patterns were modeled (*22, 23*). Shown: all-or-nothing and transmission blocking vaccine, with vaccine efficacy=90%. POL, Poland; ZAF, South Africa; CHN, China; BRA, Brazil; ZWE, Zimbabwe; ESP, Spain; IND, India; USA, United States of America; BEL, Belgium.

**Figure S10:**
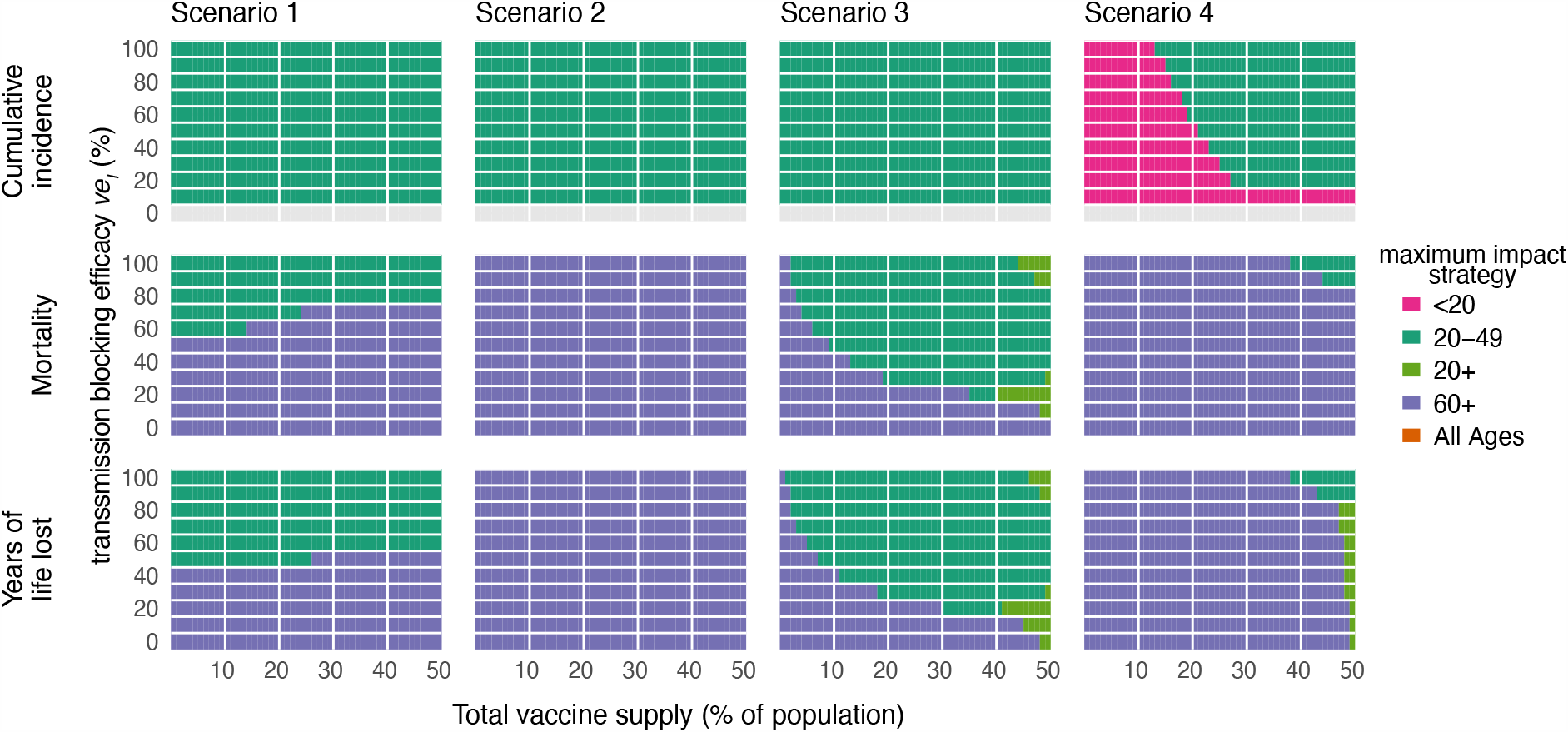
Impact of imperfect transmission-blocking effects on maximum impact strategies. Heatmaps show the prioritization strategies resulting in maximum reduction of infections (top row), mortality (middle row), and years of life lost (bottom row) across Scenario 1 (0.2% rollout/day, *R*_0_ = 1.15; left column), Scenario 2 (0.2% rollout/day, *R*_0_ = 1.5; left-middle column), Scenario 3 (pre-transmission vaccination, *R*_0_ = 1.5; right-middle column), and Scenario 4 (pre-transmission vaccination, *R*_0_ = 2.6; right column). Each heatmap shows results from simulations varying vaccine supply and the vaccine’s efficacy in blocking transmission *ve*_*I*_ as indicated. Because the bottom row of cumulative incidence plots corresponds to a vaccine with 0% efficacy to reduce transmission, the row is grey indicated no maximum impact prioritization. Shown: contact patterns and demographics of the United States (*22, 23*), for a vaccine with protective efficacy from severe disease *ve*_*P*_ = 0.9 and no efficacy for protection from infection *ve*_*S*_ = 0. See Supplementary Text for modeling details.

**Figure S11:**
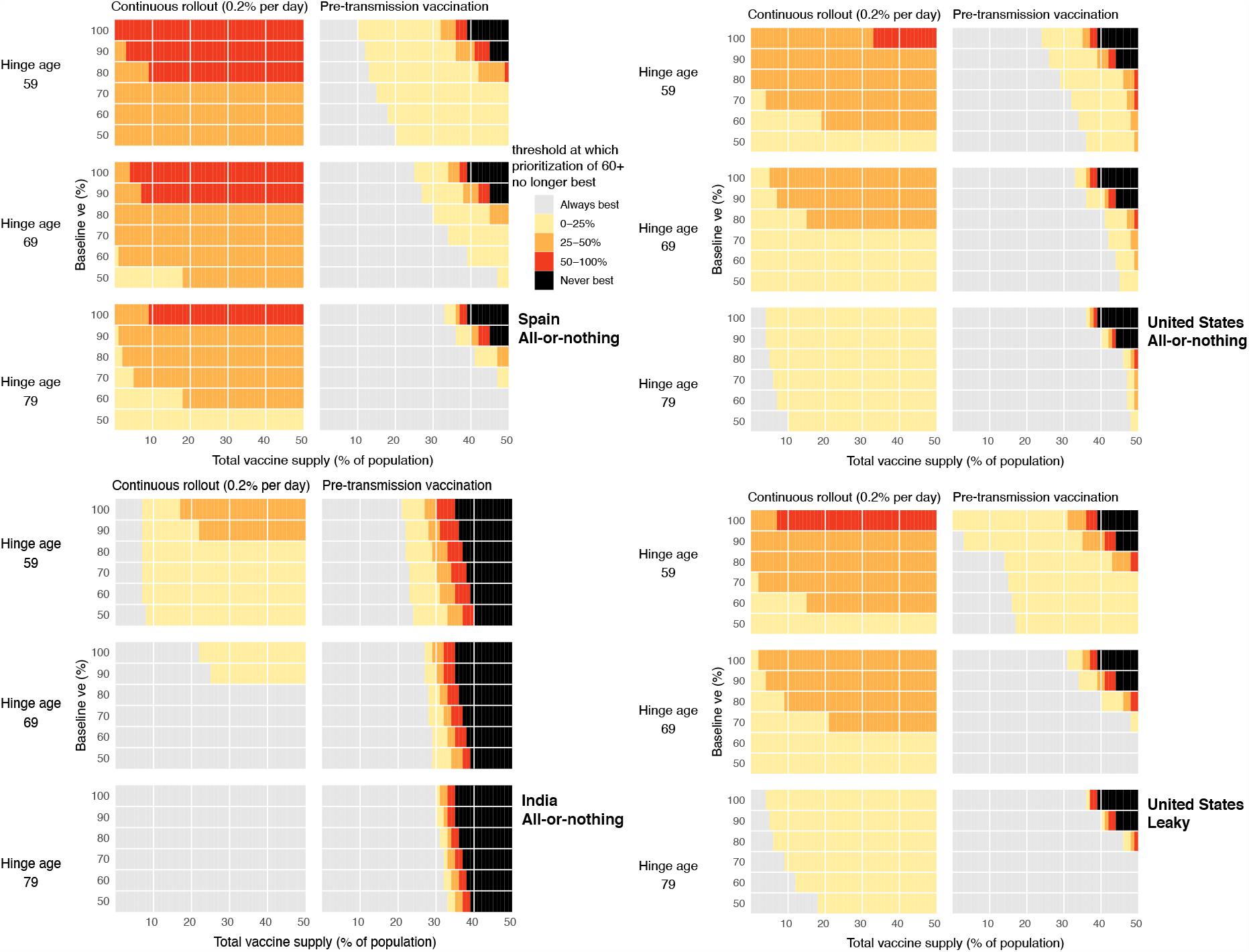
Impact of age-related decreases in vaccine efficacy on vaccine prioritization. Heatmaps show that prioritization of adults 60+ to minimize mortality remains generally robust to large decreases in vaccine efficacy among older adults. Each point shows the threshold value of vaccine efficacy among adults 80+ at which prioritizing adults 60+ is no longer the best strategy to minimize mortality, if one exists (yellow, orange, red), or indicates that none exists (grey). Parameter combinations for which mortality is never minimized by prioritization of adults 60+ are also shown (black). Panels show combinations of the age at which immunosenescence begins (hinge age), total vaccine supply, and baseline efficacy for continuous (0.2% per day, *R*_0_ = 1.5) and pre-transmission rollout scenarios with *R*_0_ = 2.6 for (A) Spain, (B) the United States, and (C) India, using an all-or-nothing vaccine model; (D) the United States, using a leaky vaccine model.

**Figure S12:**
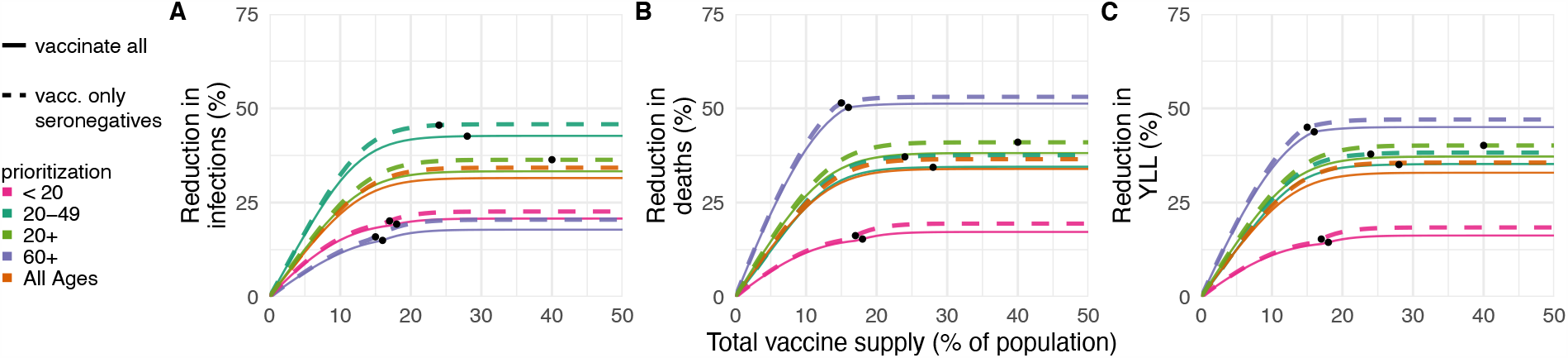
Effects of existing seropositivity on the impacts of prioritization strategies (low seroprevalence). Percent reductions in (A) infections, (B) deaths, and (C) years of life lost (YLL) for prioritization strategies when existing age-stratified seroprevalence is incorporated (July 2020 estimates for Connecticut; mean seroprevalence 3.4% (*31*)). Plots show reductions for Scenario 2 (2% rollout/day, realized *R* = 1.5) when vaccines are given to all individuals (solid lines) or to only seronegatives (dashed lines), inclusive of imperfect serotest sensitivity and specificity. Black dots indicate breakpoints at which prioritized demographic groups have been 70% vaccinated, after which vaccines are distributed without prioritization. Shown: contact patterns and demographics for the United States (*22, 23*); all-or-nothing and transmission-blocking vaccine with vaccine efficacy=90%. See Figs. 4 and S13 for moderate and higher seroprevalence examples, respectively.

**Figure S13:**
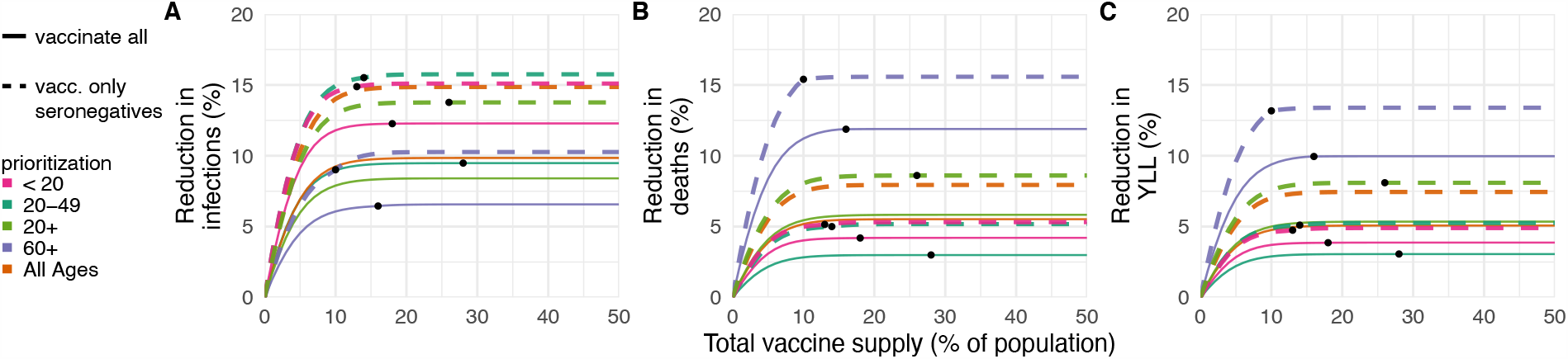
Effects of existing seropositivity on the impacts of prioritization strategies (high seroprevalence). Percent reductions in (A) infections, (B) deaths, and (C) years of life lost (YLL) for prioritization strategies when existing age-stratified seroprevalence is incorporated (model-generated; mean seroprevalence 40.1%; see Methods) and intial conditions are set to the S, E, I, and R compartment counts at the mid-outbreak time when seroprevalence reached 40.1%. Plots show reductions with 2% rollout/day, *R*_0_ = 2.6, and realized *R* = 1.43 when vaccines are given to all individuals (solid lines) or to only seronegatives (dashed lines), inclusive of imperfect serotest sensitivity and specificity. Black dots indicate breakpoints at which prioritized demographic groups have been 70% vaccinated, after which vaccines are distributed without prioritization. Shown: U.S. contact patterns and demographics (*22, 23*); all-or-nothing and transmission-blocking vaccine with vaccine efficacy = 90%. See Figs. 4 and S12 for moderate and lower seroprevalence examples, respectively.

**Figure S14:**
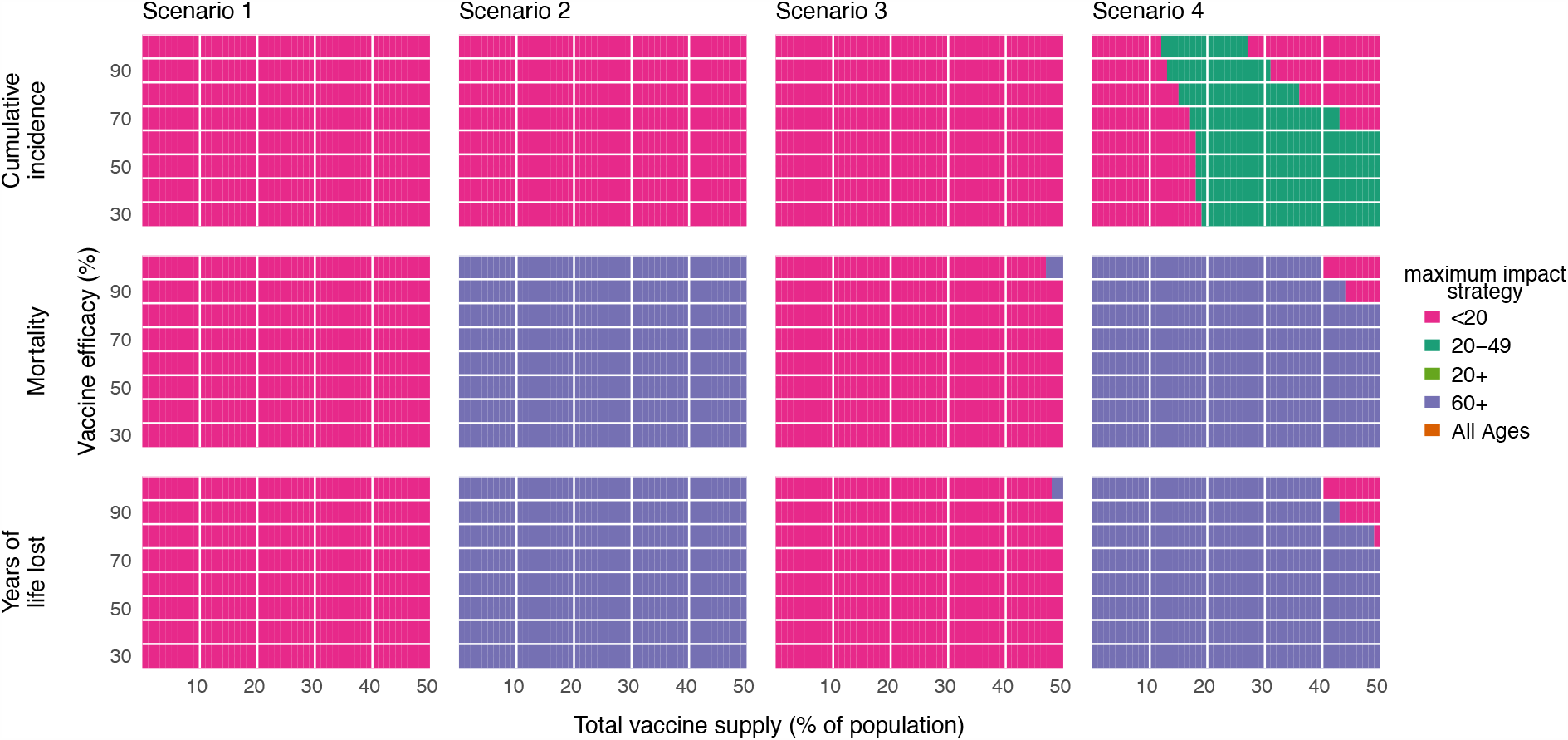
Impact of vaccine efficacy on maximum impact strategies (all-or-nothing vaccine; constant susceptibility by age). Heatmaps show the prioritization strategies resulting in maximum reduction of infections (top row), mortality (middle row), and years of life lost (bottom row) across Scenario 1 (0.2% rollout/day, *R*_0_ = 1.15; left column), Scenario 2 (0.2% rollout/day, *R*_0_ = 1.5; left-middle column), Scenario 3 (pre-transmission vaccination, *R*_0_ = 1.5; right-middle column), and Scenario 4 (pre-transmission vaccination, *R*_0_ = 2.6; right column). Each heatmap shows results from simulations varying vaccine supply and vaccine efficacy as indicated, but unlike all other simulations in this manuscript and its supplementary material, except Fig. S15, simulations use a constant susceptibility by age Shown: contact patterns and demographics of the United States (*22, 23*); all-or nothing and transmission blocking vaccine. See Fig. S15 for leaky vaccine results with constant susceptibility by age. See Fig. S5 for all-or-nothing vaccine results with varying susceptibility by age.

**Figure S15:**
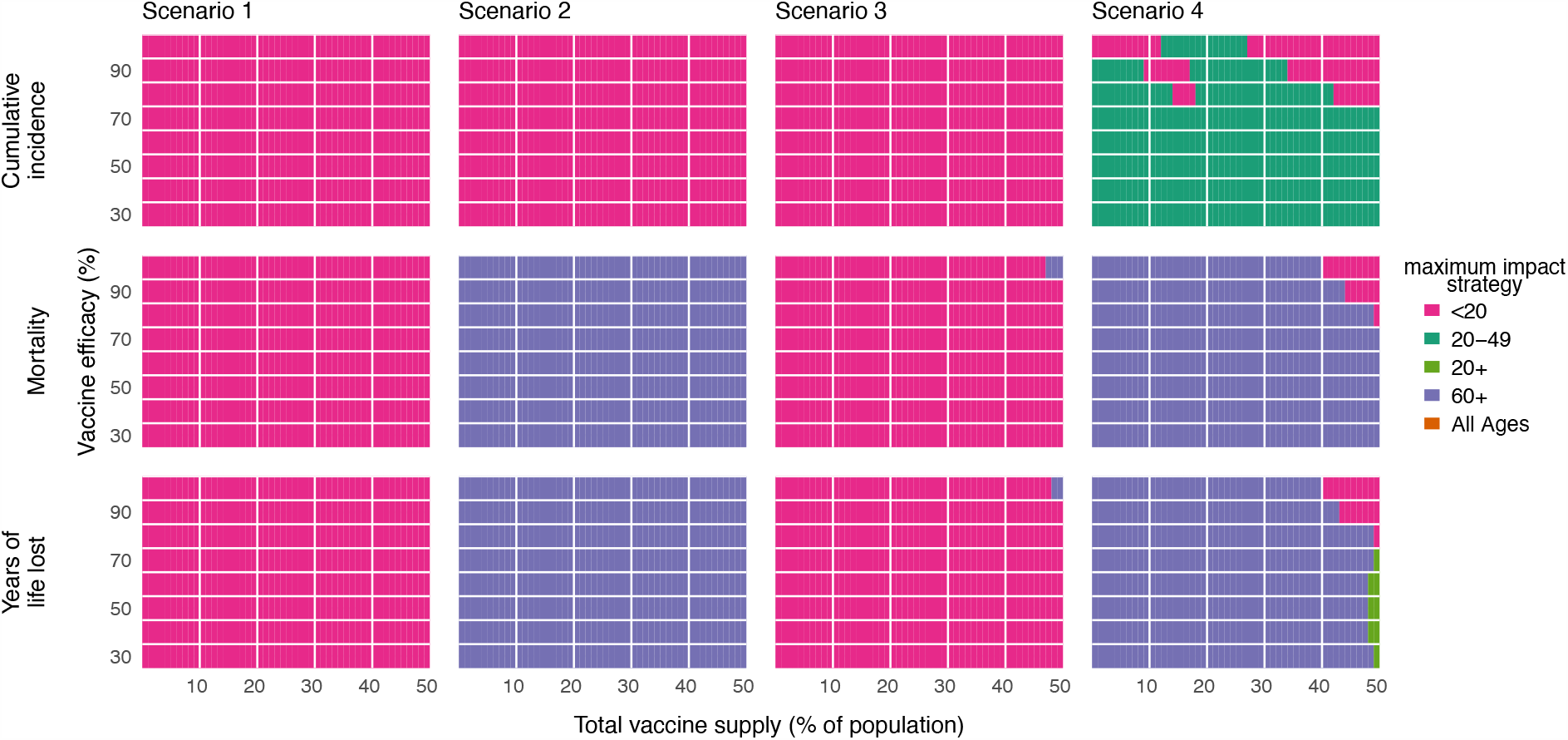
Impact of vaccine efficacy on maximum impact strategies (leaky vaccine; constant susceptibility by age). Heatmaps show the prioritization strategies resulting in maximum reduction of infections (top row), mortality (middle row), and years of life lost (bottom row) across Scenario 1 (0.2% rollout/day, *R*_0_ = 1.15; left column), Scenario 2 (0.2% rollout/day, *R*_0_ = 1.5; left-middle column), Scenario 3 (pre-transmission vaccination, *R*_0_ = 1.5; right-middle column), and Scenario 4 (pre-transmission vaccination, *R*_0_ = 2.6; right column). Each heatmap shows results from simulations varying vaccine supply and vaccine efficacy as indicated, but unlike all other simulations in this manuscript and its supplementary material, except Fig. S14, simulations use a constant susceptibility by age. Shown: contact patterns and demographics of the United States (*22, 23*); leaky and transmission blocking vaccine. See Fig. S14 for all-or-nothing vaccine results with constant susceptibility by age. See Fig. S7 for leaky vaccine results with varying susceptibility by age.

## Supplementary Tables

**Table S1:**
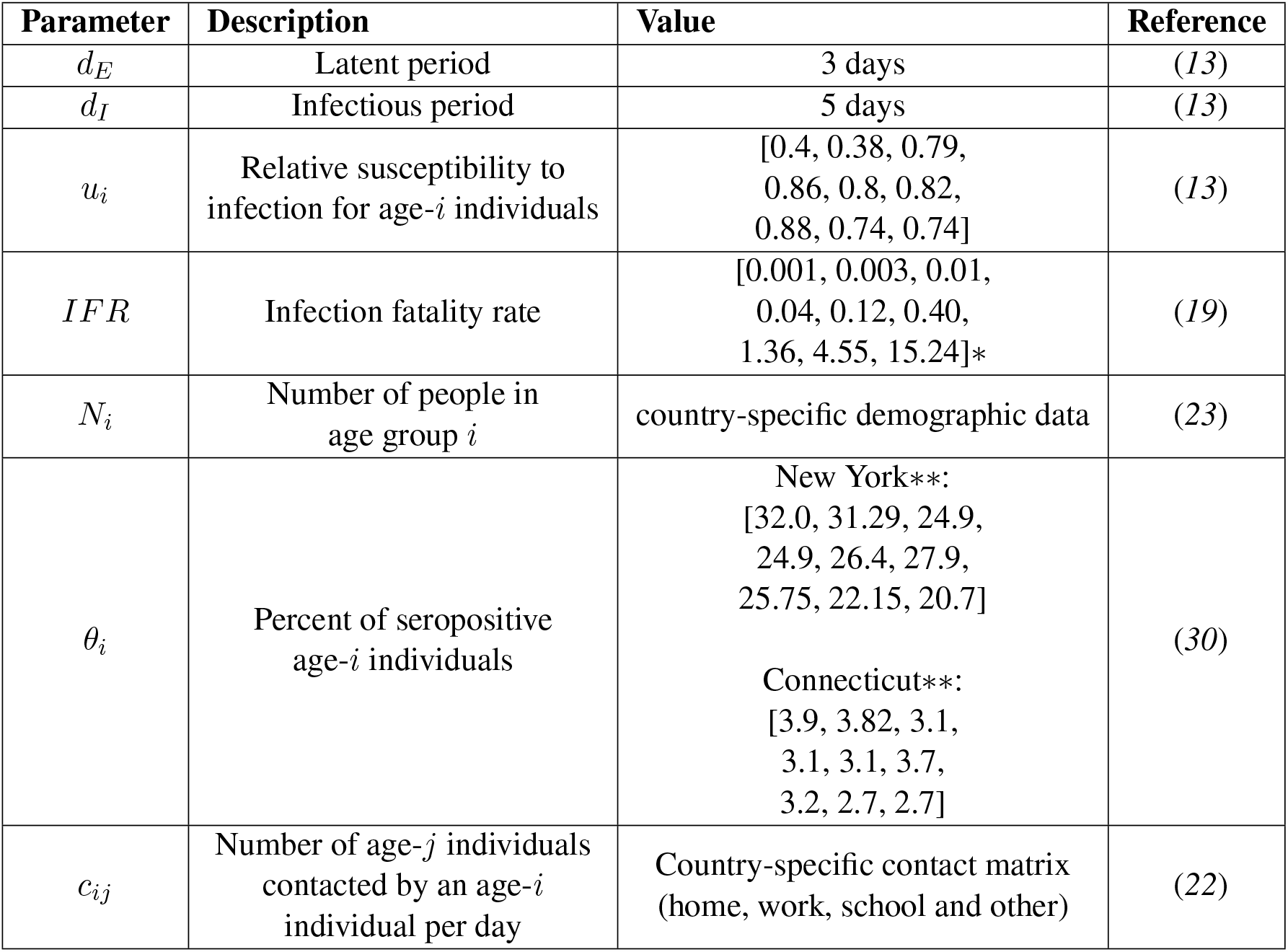
Summary of parameters used in modeling and simulation. *\**To relate the IFR metaregression result from (*19*), *log*_10_(IFR) = *-*3.27 + 0.0524 *\** IFR age to age bins by decade, we computed IFR_*i*_ = (Σ _*j*_ IFR_*j*_)*/*10 where *j* are the ages (in years) corresponding to bin *i*. *\*\**To relate NYC (*30*) and Connecticut (*31*) seroprevalence estimates to age bins by decade, we computed *θ*_*i*_ = (Σ _*j*_*j θ*_*j*_)*/*10 where *j* are the ages (in years) corresponding to bin *i*.

**Table S2:** Initial conditions for simulations. Scenario initial conditions (ICs) for age group *i* are reported for scenarios considered in this manuscript. NA, Not Applicable; *se*, sensitivity; *sp*, specificity; *vh*, vaccine hesitant fraction. *S* = *N*_*i*_ *-I*_*i*_(0) *-I*_*x,i*_ (0) *-θ*_*i*_*N*_*i*_. The proportion of each age group that is vaccine hesitant is denoted *vh* and not considered for vaccination. Vaccination rollout *α*_*i*_ = *n*_vax,*i*_*/* [(*S*_*i*_ + *E*_*i*_)*sp* + *R*_*i*_(1 − *se*)]. Note that modeling without targeting of seronegatives via serological testing is equivalent to setting *sp* = 1 and *se* = 0. * ICs for all other compartments not explicitly specified are 0.

| IC* | Pre-transmission rollout,<br>all-or-nothing $ve$ | Pre-transmission rollout,<br>leaky $ve$ | Continuous rollout |
| --- | --- | --- | --- |
| $I_i(0)$ | 1 | | $0.0025 N_i(1 - vh)$ |
| $I_{x,i}(0)$ | 1 | | $0.0025 N_i(vh)$ |
| $E_i(0)$ | 0 | | $0.0025 N_i(1 - vh)$ |
| $E_{x,i}(0)$ | 0 | | $0.0025 N_i(vh)$ |
| $R_i(0)$ | $\theta_i N_i(1 - vh) - \alpha_i \theta_i N_i(1 - vh)$ | | $\theta_i N_i(1 - vh)$ |
| $R_{v,i}(0)$ | $\alpha_i \theta_i N_i(1 - vh)(1 - se)$ | | 0 |
| $R_{x,i}(0)$ | $\theta_i N_i(vh) + \alpha_i \theta_i N_i(1 - vh)se$ | | $\theta_i N_i(vh)$ |
| $S_i(0)$ | $S(1 - vh) - \alpha_i S(1 - vh)$ | | $(N_i - 0.005N_i - \theta_i N_i)(1 - vh)$ |
| $S_{v,i}(0)$ | NA | $\alpha_i S(1 - vh)(sp)$ | 0 if leaky $ve$,<br>else NA |
| $V_i(0)$ | $\alpha_i S(1 - vh)ve(sp)$ | NA | 0 if all-or-nothing $ve$,<br>else NA |
| $S_{x,i}(0)$ | $S(vh) +$<br>$\alpha_i S(1 - vh)(1 - sp) -$<br>$\alpha_i S(1 - vh)(1 - ve)(sp)$ | $S(vh) +$<br>$\alpha_i S(1 - vh)(1 - sp)$ | $(N_i - 0.005N_i - \theta_i N_i)(vh)$ |

